# A novel COVID-19 epidemiological model with explicit susceptible and asymptomatic isolation compartments reveals unexpected consequences of timing social distancing

**DOI:** 10.1101/2020.05.11.20098335

**Authors:** Jana L. Gevertz, James M. Greene, Cynthia Sanchez-Tapia, Eduardo D. Sontag

## Abstract

Motivated by the current COVID-19 epidemic, this work introduces an epidemiological model in which separate compartments are used for susceptible and asymptomatic “socially distant” populations. Distancing directives are represented by rates of flow into these compartments, as well as by a reduction in contacts that lessens disease transmission. The dynamical behavior of this system is analyzed, under various different rate control strategies, and the sensitivity of the basic reproduction number to various parameters is studied. One of the striking features of this model is the existence of a critical implementation delay (“CID”) in issuing separation mandates: while a delay of about two weeks does not have an appreciable effect on the peak number of infections, issuing mandates even slightly after this critical time results in a far greater incidence of infection. Thus, there is a nontrivial but tight “window of opportunity” for commencing social distancing in order to meet the capacity of healthcare resources. However, if one wants to also delay the timing of peak infections –so as to take advantage of potential new therapies and vaccines– action must be taken much faster than the CID. Different relaxation strategies are also simulated, with surprising results. Periodic relaxation policies suggest a schedule which may significantly inhibit peak infective load, but that this schedule is very sensitive to parameter values and the schedule’s frequency. Furthermore, we considered the impact of steadily reducing social distancing measures over time. We find that a too-sudden reopening of society may negate the progress achieved under initial distancing guidelines, but the negative effects can be mitigated if the relaxation strategy is carefully designed.

## 1 Introduction

Early 2020 saw the start of the coronavirus disease 2019 (COVID-19) pandemic, which is caused by severe acute respiratory syndrome coronavirus 2 (SARS-CoV-2). Current COVID-19 policy is being largely influenced by mathematical models [1, 2, 3, 4, 5, 6, 7, 8, 9]. Some of these are classic epidemiological ordinary differential equations (ODE) models. Such models are suitable for describing initial stages of an infection in a single city, as well as for describing late stages at which transportation effects are small in comparison to community spread. Besides being simpler to analyze mathematically, ODE models are also a component of more complex network simulations that incorporate interacting populations linked by transportation networks as well as social, educational, and workplace hubs. The work described here is in the spirit of the former, ODE models.

We have developed and analyzed a variation of the classic epidemiological SIR model which incorporates separate “compartments” for “socially distanced” healthy and asymptomatic (but infected) populations, as well as for infected (symptomatic) populations. There have been many models proposed in the literature to deal with “quarantined” populations, see for example [10, 11], but, to the best of our knowledge, no models in which susceptible populations are split into non-distanced and distanced sub-classes in such a way that the rates of flow between these are viewed as control variables. Indeed, key to our model are parameters that reflect the rate at which individuals become “socially distant” and the rate at which individuals return to the “non-distanced” category. As examples, the latter might represent a “frustration” with isolation rules, or a personal need to reduce the economic impact of social distancing. The former can be in principle manipulated by government intervention, through the strength of persuasion, and law enforcement.

How do outcomes depend on such interventions? How does one trade-off various types of other interventions (for example vaccination, which would affect transmissibility, or curfew rules) against each other? Our modeling work aims to provide a framework to rigorously formulate and answer such questions.

We will view the rate at which individuals respond to mandates as a control variable, and analyze the impact of different control policies on the course of an epidemic. *A novel aspect of our model lies in the distinction that we make between rate control and the decrease in contacts between infected and susceptible individuals due to distancing*. We call this latter reduction in transmission the *contact rescaling factor (CoRF)*. One can interpret the CoRF value as reflecting the effectiveness of social distancing. This number is a function of the stringency of rules (stay at home except for shopping and emergencies, wash hands frequently, wear masks, stay 6 ft. apart, etc.). Some authors consider tuning what we call the CoRF as the control “knob” used by authorities, e.g. [12, 13, 14, 15, 16]. Our focus is, instead, on rate control, which has not been sufficiently explored. Indeed, the objective of our model is to make it possible to formally consider rate control. In future work, we will study the combination of rate and CoRF control.

In particular, we used our model to answer questions about the dynamics of the disease, and about the value of the basic reproduction number, *R*_0_, which characterizes the initial rise in infections. We rigorously demonstrate, without simulations, that at sufficiently early stages of the pandemic when there is little immunity in the population, a (possibly unrealistic) quick implementation of social distancing is required in order for *R*_0_ < 1. While it is easy to interpret this as a hopeless situation, what this actually says is that an initially headline-grabbing infection will begin to move through the population. However, as time progresses, we show that social distancing can push *R*_0_ to a value less than 1.

This conclusion about the impact of social distancing at different stages on the pandemic is dependent on the parameterization of the model. As many of the parameter values are still uncertain, we also explored how *R*_0_ depends on a combination of a single model parameter and the social distancing rate parameter. One major uncertainty surrounding COVID-19 is the fraction of individuals who get infected but never develop symptoms. We find that *R*_0_ is sensitive to this symptomatic fraction, demonstrating the importance of getting a confident measurement on this value before quantitative model predictions can be trusted.

Another major unknown is how infective asymptomatic individuals are. We find that if asymptomatics are not very contagious, and if infected individuals automatically self-isolate, then *R*_0_ is not greatly influenced by social distancing measures. However, if asymptomatics are sufficiently infective, there is a much stronger impact of social distancing on *R*_0_. That said, this conclusion depends on the assumption that social distancing reduces the transmission rate of the disease by the value of the CoRF. Therefore, varying this parameter allows us to quantify how the nature of social distancing measures impacts *R*_0_. If this parameter is very small, meaning one significantly down-scales their contacts (that is, the stay-at-home directives are extreme), very rapid implementation of social distancing is not required. On the other hand, if the directives are not as severe and CoRF is larger (meaning the number of contacts is scaled down less significantly), social distancing will not result in *R*_0_ < 1 and we can still expect disease spread despite social distancing.

We also used our model to explore how the timing of when social distancing is enacted influences the spread of the disease through a population. One of the most striking predictions is that a moderate delay in establishing social distancing guidelines, which we term a *critical implementation delay (CID)*, does not appreciably increase the *peak number* of infected individuals. Keeping this number low (“flattening the curve”) is desirable in order to prevent strain on health providers and hospital resources. The existence of a CID means that authorities can take some time to plan for guidelines and announce a closure plan. Another important feature of the CID is that even a few days delay in implementation beyond the CID can have highly adverse consequences. Once passed, there will be many more (over ten times) the number of sick people in the population at its worst moment. *With our parameters*, the CID is roughly two weeks.

That said, there are good reasons both for and against taking advantage of the CID. We find that implementing *even faster* than the CID time results in a major postponement of the peak *time* for infections. For example, a delay of 15 days has a peak of infection early on in the epidemic, occurring at about 50 days. However, initiating 10 days earlier delays the peak to almost one year, a huge difference in timing. Such a postponement provides more opportunities to develop vaccines and treatments, and hence can be seen as highly desirable. On the other hand, extended waiting may also create anxiety. This trade-off must be carefully considered by policy makers and society at large.

Related to timing, there has been interest in periodically relaxing distancing guidelines to allow for limited economic activity. For example, businesses may be allowed to operate normally for one week, while the ensuing week is restricted to remote operation (or being fully closed, if remote work is not feasible). This two week “periodic” schedule is then continued either for a fixed period of time, or indefinitely (e.g. the discovery of a vaccine, evidence that sufficient herd immunity has been obtained, etc.) Using our model with estimated parameters, we simulate such schedules for a variety of periods, ranging from days to months of sanctioned activity. Our results are quite counter-intuitive, and suggest that there might be a pulsing period that significantly inhibits the infection dynamics (a 17 day “on/off” schedule with our parameters). However, this schedule is exceptionally sensitive to parameter values and timing, *so that extreme caution must be taken when designing guidelines that fully relax social distancing, even temporarily*. Furthermore, for some strategies near the optimal 17-day cycle, a subsequent increase in infected individuals may occur *after* an initial flattening. Thus, even if a region observes a short-term improvement, the *worst may still be yet to come*.

Other forms of relaxation relate to gradual easing, as opposed to periods of “normal activity.” Of course, the rate of easing (e.g. how many people are allowed in a grocery store or in an office) is of great interest, both economically and psychologically. We numerically investigate how the rate of easing social distancing guidelines affects outbreak dynamics, and show that relaxing too quickly will only delay, *but not suppress*, the peak magnitude of symptomatic individuals. However, a more gradual relaxation schedule will both delay onset and “flatten the curve,” while producing a largely immune population after a fixed policy window (again, assuming recovery corresponds to immunity, which is still an open question as of this writing). Hence the rate of relaxation is an important factor in mitigating the severity of the current pandemic. Similarly, the rate of relaxation *during flattening* is important to prevent a “second wave” of infected individuals. As governments develop and implement plans to ease social distancing, carefully considering the rate of relaxation is extremely important from a policy perspective, so that countries and states do not undo the benefits of their strict distancing policies by lifting guidelines too rapidly. For example, *in our model*, a very rapid relaxation schedule results in a second wave with a larger peak symptomatic proportion than originally experienced (over 27%, compared to original peak of 3.2%). However, relaxing more gradually once the peak has been obtained prevents a second outbreak, and allows a sustainable approach to herd immunity.

We close this introduction with the following quote:

> “I have skepticism about models [of COVID-19], and they are only as good as the assumptions you put into them, but they are not completely misleading. They are telling you something that is a reality, that when you have mitigation that is containing something, and unless it is down, in the right direction, and you pull back prematurely, you are going to get a rebound of cases.”
>
> Dr. Anthony Fauci, Director, National Institute of Allergy and Infectious Diseases, United States; on CNN, 05 May 2020

It bears emphasizing: ours is one model, with one set of assumptions. We do not in any way believe that the quantitative predictions of our (or of any other) model of COVID-19 can be accurate, as so much is still unknown about this disease. However, as in the statistician George Box’s aphorism “All models are wrong, but some are useful”, the correct question is not if the model is “true” but rather if it is “illuminating and useful.”

## 2 Models

The SIR model proposed by W.O. Kermack and A.G. McKendrick in 1927 [17] has been applied in many ways over the last century to study infectious diseases, and recently has been extended to study COVID-19. For example, a recent model for COVID-19, called the SIDARTHE model [12], partitions individuals as susceptibles, asymptomatic and undetected infected, asymptomatic detected, symptomatic undetected, symptomatic detected, detected with life-threatening symptoms, recovered, and deceased. There are also several papers that deal with timing of interventions as well as periodic strategies to prevent the spread of epidemics, modeled through periodic vaccination [18] or through the periodic or other switching of the infectivity parameter in SIR and related models [12, 14, 15, 16].

In this work we propose a different extension of the SIR model, one that includes socially distanced (labeled with a *D* sub-index) and non-socially distanced (labeled with an *N* sub-index) classes for susceptible (*S_D_* and *S_N_*), asymptomatic (*A_D_* and *A_N_*), and symptomatic (*I_D_* and *I_N_*) individuals. Class *R* refers to “Recovered” who are presumed to have developed at least temporary immunity. More details about the interpretation of each variable together with the meaning of the variables and parameters used in this model can be found at Table 1. Next, we explain the dynamics of our model (please refer to Fig. 1 for a graphical explanation).

1. A socially distanced susceptible individual (*S_D_*) may become infected with rate:

- ***ϵ_S_β_A_A_N_*** when in contact with a non-socially distanced asymptomatic individual. Here, *β_A_* is the transmission rate between an asymptomatic non-socially distanced individual and a non-socially distanced susceptible; and the term *ϵ_S_* accounts for the reduction of infectivity by socially distancing the susceptible. We call *ϵ_S_* a contact rescaling factor (CoRF).
- ***ϵ_S_ϵ_A_β_A_A_D_*** when in contact with a socially distanced asymptomatic individual. The term *ϵ_S_ϵ_A_* refers to the reduction of infectivity by socially distancing both the susceptible and the asymptomatic individuals.
- ***ϵ_S_β_I_I_N_*** when in contact with a non-socially distanced symptomatic individual. The term *β_I_* denotes the transmission rate between non-socially distant symptomatic and non-socially distanced susceptible individuals.
- ***ϵ_S_ϵ_I_β_I_I_D_*** when in contact with a non-socially distanced symptomatic individual. The term *ϵ_S_ϵ_I_* denotes the reduction of infectivity by socially distancing both the susceptible and the symptomatic individuals. We expect that socially distanced symptomatic individuals are still capable of transmitting infections, be it through contact with hospital personnel or caregivers, or the pressure to work despite being sick.
2. Similarly, a non-socially distanced susceptible individual (*S_N_*) may become infected with rate:

- ***β_A_A_N_*** when in contact with a non-socially distanced asymptomatic individual.
- ***ϵ_A_β_A_A_D_*** when in contact with a socially distanced asymptomatic individual.
- ***β_I_I_N_*** when in contact with a non-socially distanced symptomatic individual.
- ***ϵ_I_β_I_I_D_*** when in contact with a socially distanced symptomatic individual.
3. If a susceptible individual that has been social distancing (an individual in class *S_D_*) gets infected, they will continue social distancing (will transfer to class *A_D_*); and a non-social distanced individual will continue non-social distancing right after getting infected (will transition from the *S_N_* to the *A_N_* class).
4. Susceptible individuals transition from social distancing to non-social distancing behavior with rate *h*_1_. Likewise for asymptomatic individuals.
5. Susceptible individuals transition from non-social distancing to social distancing behavior with rate *h*_2_. Likewise for asymptomatic individuals.
6. After the incubation period, an asymptomatic individual may or may not become symptomatic. Thus, an individual may transition from the asymptomatic class into the symptomatic class, or directly to the recovered class. The parameter f represents the fraction of the asymptomatic individuals that transition into the symptomatic class. Thus (1 − *f*) is the fraction of individuals who are asymptomatic and transition directly to the recovered group.
7. The transition rate out of asymptomatic, *γ_AI_*, is independent of whether one was socially distancing or not.
8. A fraction *p* of non-socially distanced asymptomatic start social distancing after becoming symptomatic. Thus, (1 − *p*) is the fraction of non-socially distanced asymptomatic individuals that remain non-social distancing after becoming symptomatic.
9. A social distancing asymptomatic that becomes symptomatic remains socially distancing (transfers from *A_D_* into *I_D_*).
10. If an individual becomes symptomatic, they will either recover (transfer to the *R* class with rate *γ_IR_*) or die with rate *δ*.
11. Recovery assumes that the individual will acquire temporary immunity.
12. Recovered individuals lose immunity at a rate *ρ*.
13. A fraction *q* of recovered individuals who lost immunity remain socially distanced, and a fraction (1 − *q*) will stop social distancing.

**Figure 1:**
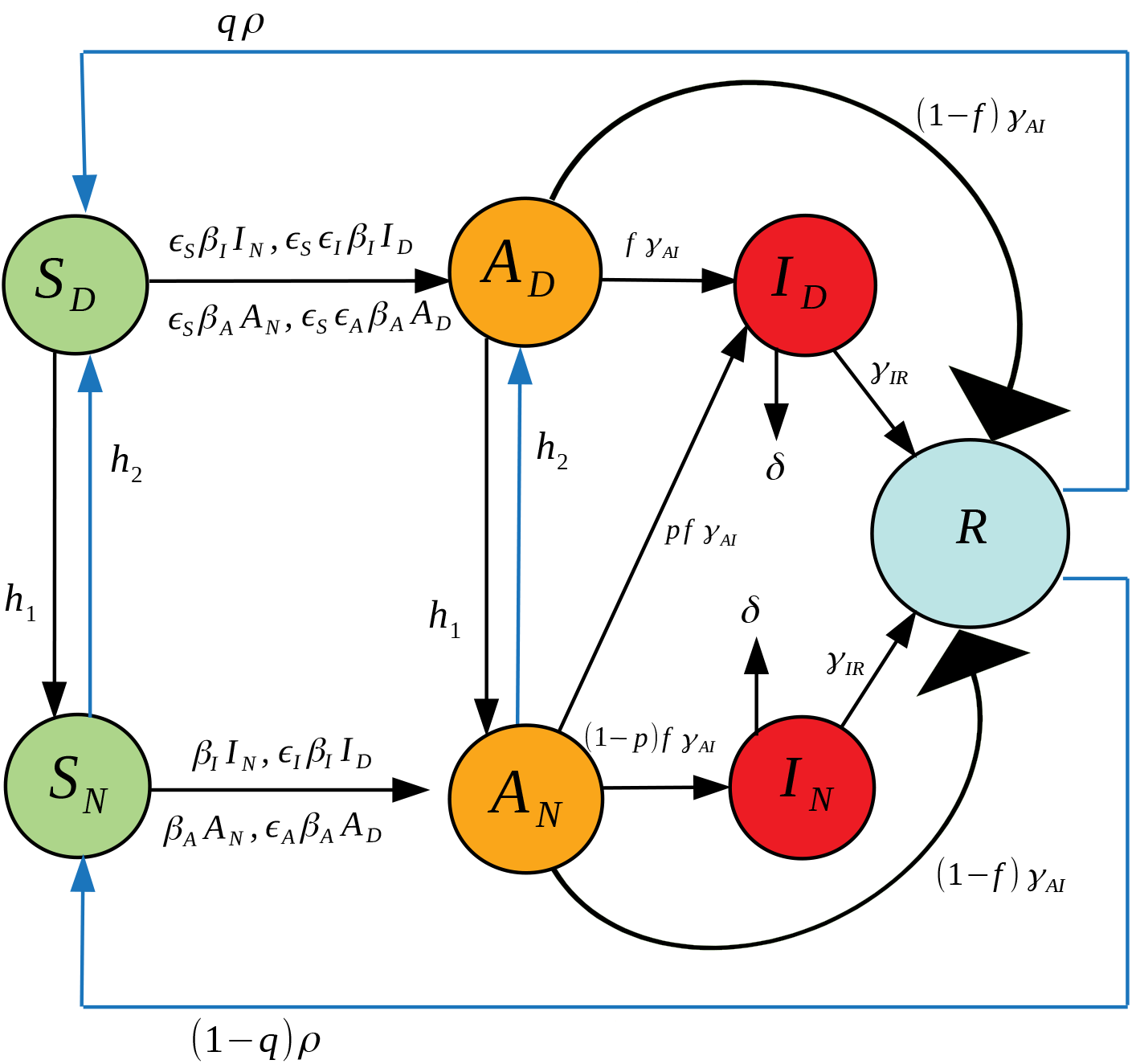
Illustration of the seven compartment SIR model in equations (1)-(7).

**Table 1:** List of all the variables and parameters used in both models: the seven-compartment model in equation (1)-(7), and the six-compartment model in equations (8)-(13).

| Symbol | Interpretation |
| --- | --- |
| $S_N$ | Fraction of population that are non-socially distant susceptible. |
| $S_D$ | Fraction of population that are socially distant susceptible. |
| $A_N$ | Fraction of population that are non-socially distant asymptomatics. |
| $A_D$ | Fraction of population that are socially distant asymptomatics. |
| $I_N$ | Fraction of population that are non-socially distant symptomatics. |
| $I_D$ | Fraction of population that are socially distant symptomatics. |
| $R$ | Fraction of population that are recovered and presumed to have developed at least temporary immunity. |
| $\epsilon_S$ | CoRF: Effect of socially distancing susceptibles on disease transmission. |
| $\epsilon_A$ | CoRF: Effect of socially distancing asymptomatics on disease transmission. |
| $\epsilon_I$ | CoRF: Effect of socially distancing symptomatic infectives on disease transmission. |
| $\beta_A$ | Transmission rate between asymptomatic non-socially distanced individual and non-socially distanced susceptible. |
| $\beta_I$ | Transmission rate between non-socially distanced symptomatic and non-socially distanced susceptible. |
| $\delta$ | Mortality rate of disease. |
| $f$ | Fraction of individuals who become symptomatic (as opposed to never showing symptoms and recovering). |
| $h_1$ | Return to socializing rate. |
| $h_2$ | Rate of social distancing (control). |
| $\gamma_{AI}$ | Rate of transition out of the asymptomatic class. |
| $\gamma_{IR}$ | Rate of recovery. |
| $p$ | Fraction of non-socially distanced asymptomatics who socially distance upon showing symptoms. |
| $q$ | Fraction of recovered individuals who lose immunity but continue social distancing. |
| $\rho$ | Rate at which recovered individuals lose immunity and become susceptible again. |

The differential equation system representing this seven-compartment model is as follows:

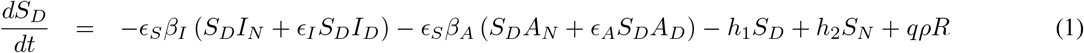

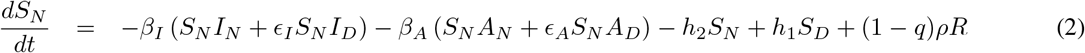

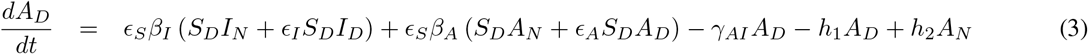

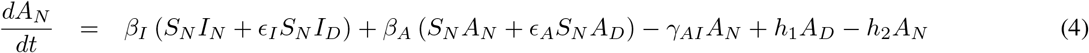

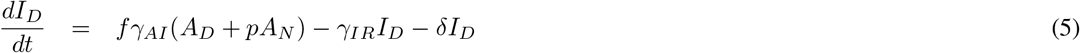

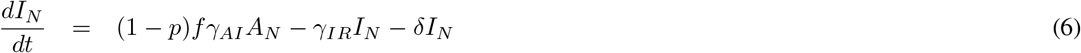

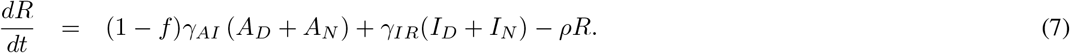

Although very little is known about immunity in regards to COVID-19, it is known that for other types of coronaviruses such as the severe acute respiratory syndrome (SARS), antibodies are maintained for an average of two years [19, 20, 21]. At present, pharmaceutical companies around the world are working to develop a vaccine for COVID-19, and it is hoped that one will be widely deployed in less than two years. For this reason, we are currently interested in understanding the dynamics that will occur during the waiting period for a vaccine. It seems reasonable then, under the assumption that a recovered individual will acquire immunity for an average period of two years, to start by studying the simplified model where immunity is not temporary. Further, given the widespread understanding of the contagious nature of SARS-CoV-2, it is also reasonable to assume that symptomatic individuals self-isolate.

### 2.1 A simplified version (A six compartment SIR Model)

In this simplified model we assume permanent immunity for the recovered class, and that all symptomatics (*I_N_* and *I_D_*) can be merged into just one class *I* (see Fig. 2). The differential equation system representing this model is given below:

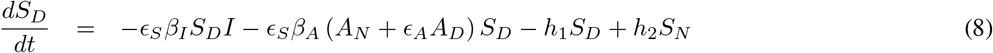

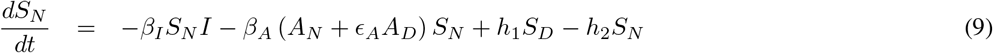

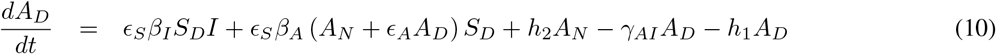

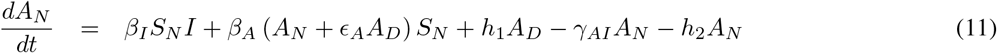

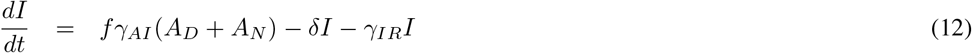

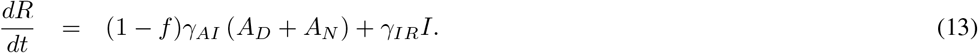

**Figure 2:**
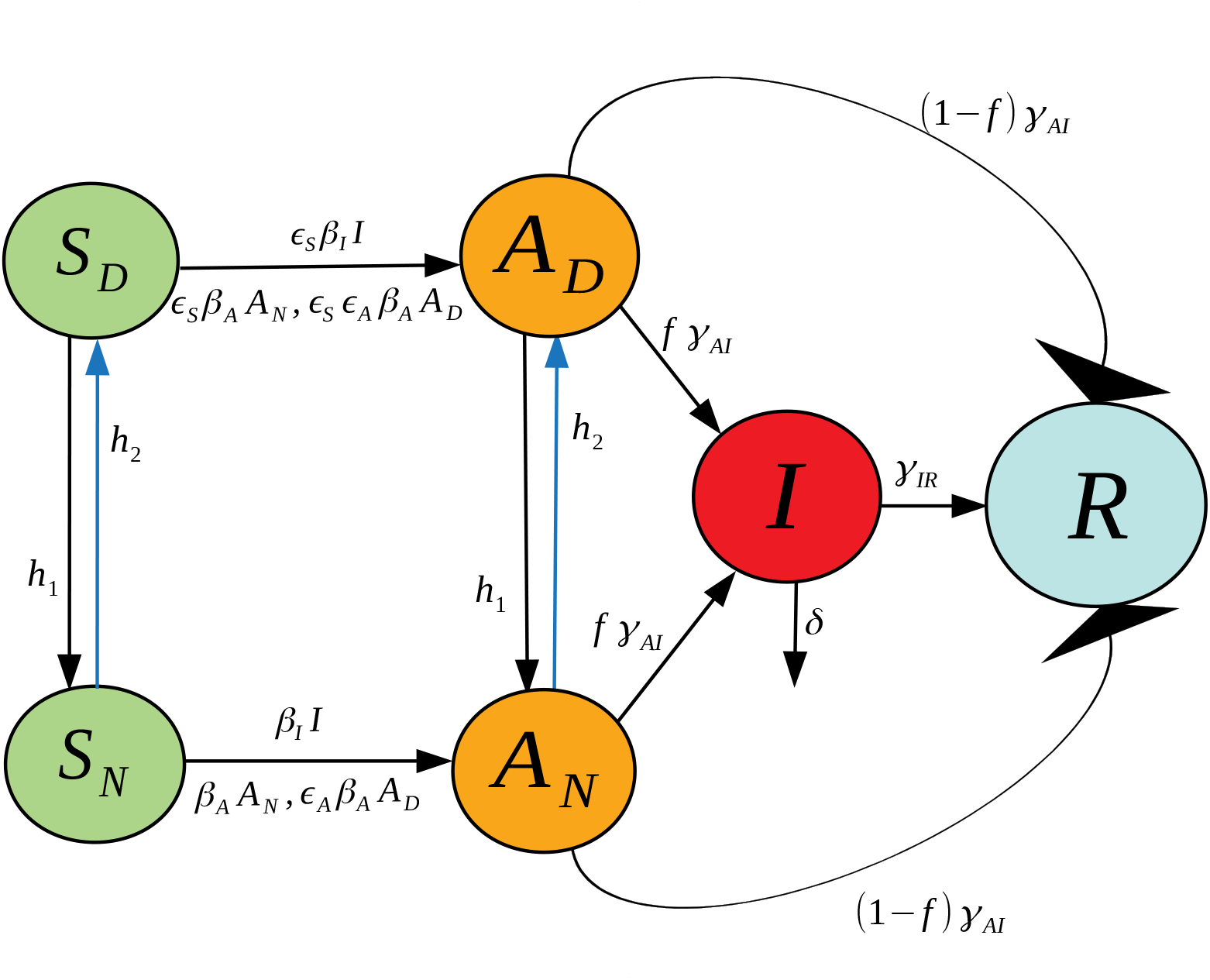
Illustration of the six compartment SIR model in equations (8)-(13).

### 2.2 Parameter estimation from currently available data

We first note that the variables in our model system (8)- (13) should be interpreted in terms of fractions of the population, and not as absolute population numbers. Of course, a direct translation is possible by fitting to a region of interest, and multiplying by the total population size at the time of disease outbreak. Note that since the initial conditions are all non-negative and satisfy

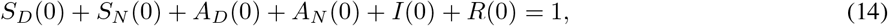

then all variables remain in the interval [0,1] for future times *t* > 0, and can hence be interpreted as a fraction of the initial population. Note that if *δ* > 0 (a strictly positive death rate) and (14) is satisfied, the total population fraction *N* defined by

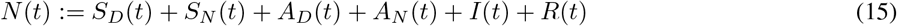

will satisfy

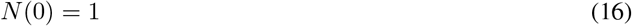

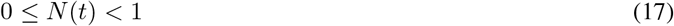

for all *t* > 0. In fact, the difference 1 − *N*(*t*) measures the fraction of deaths of the initial population by time *t*. Births are ignored, since we consider time-scales on the order of 1 year, and newborns are not significant contributors to the susceptible populations.

It is easy to verify (see Appendix C) that all the infective populations, *A_N_* (*t*), *A_D_* (*t*), and *I*(*t*) converge to zero as *t* → ∞.

One contribution of this work is to make the distinction between rate control and decrease in contacts due to social distancing. Hence, we need to explicitly define rate control (*h*_1_ and *h*_2_) in our model. To this end, recall that *h*_1_is interpreted as a socializing rate, while *h*_2_ is a controlled level of social distancing. Intuitively, we expect that increasing social distancing guidelines will at the same time inhibit individuals from socializing. That is, *h*_1_ and *h*_2_ are not independent, but are rather inversely correlated to one another. To make this mathematically precise, we define

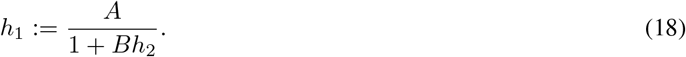

That is, increasing distancing mandates (*h*_2_) at the same time decreases the rate at which individuals socialize (*h*_1_). Other functional relationships are possible, but for the remainder of this work we fix *h*_1_ as in equation (18). Furthermore, we fix

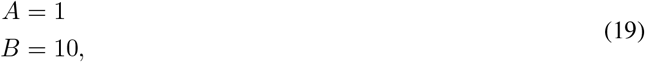

so that

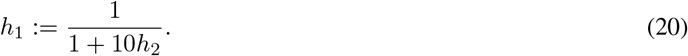

It is difficult to estimate such rates directly, as they correspond to sociological responses to unprecedented self-isolation guidelines. However, our rationale is as follows. Consider first a policy such that *h*_2_ = 1 per day, which implies that (interpreting the ODE system as the expected value of the corresponding Poisson process) that the average time to socially distance is

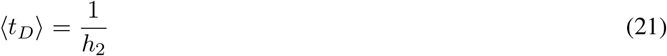

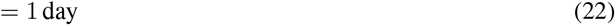

Assuming that the population is initially non-distanced, so that *S_N_* (0) ≈ 1,*S_D_* (0) ≈ 0, and ignoring the infection dynamics over a period of 1 day, we have the estimate

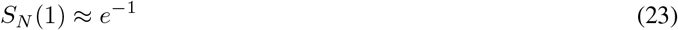

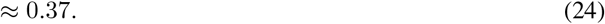

In the above, we ignored transitions from *S_D_* into *S_N_*, as *S_D_* is assumed small. Hence, after 1 day, approximately 63% of the population socially distances. Equation (20) then yields 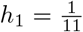, so that of the (assumed small) socially distanced population,

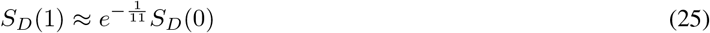

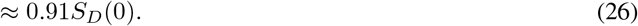

That is, about 9% of the population disobeys the distancing mandate per day. Similarly, when *h*_2_ = 0, i.e. there are no social distancing directives, equation (20) yields *h*_1_ = 1, so that

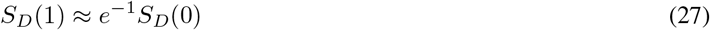

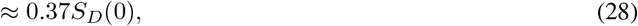

i.e. about 63% of the population re-socializes in 1 day; others may be too scared, or simply not prone to leave their house every day. The above reasoning seems at least reasonable to the authors. Of course, the focus on the subsequent analysis will not be on precise predictions, but rather general phenomenon, which are robust to parameter values. This should be considered for the remainder of this section (and the remainder of the work) as we discuss other estimates.

We can also interpret *h*_1_ and *h*_2_ in terms of the equilibrium fractions of socially distanced individuals. Indeed, in the absence of any infection (assuming no recovery has yet taken place), we have that the equilibrium fractions of *S_N_* and *S_D_* are given by

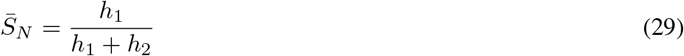

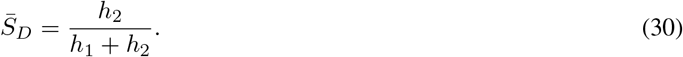

If as above, *h*_2_ = 1, then 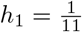, and we have that

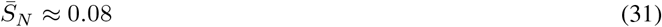

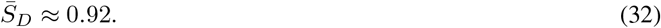

Hence, in the long-term, with very strict distancing guidelines, approximately 92% of the susceptible population will distance, while 8% do not. This also seems reasonable with very strict mandates, as of course some jobs remain essential and hence not all workers can become isolated (nurses, doctors, grocery workers, first responders, etc.).

Recall that *fγ_AI_* is the transition rate from the asymptomatic (but infected) populations *A_D_* and *A_N_*. Again interpreting as the expected value of a Poisson process, we can relate *fγ_AI_* to the expected time until asymptomatic individuals shows symptoms

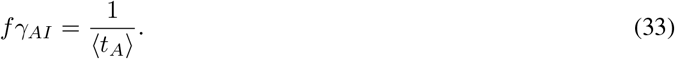

That is, *fγ_AI_* is inversely proportional the incubation period of the disease. There are different estimates of this incubation period. The original analysis based on 88 confirmed cases in Chinese provinces outside Wuhan, using data on known travel to and from Wuhan, gave an estimate of 6.4 days [22]. Later estimates of community spread have been closer to 5.1 days, with a 95% confidence interval of 4.1 to 7.0 days [23, 24]. We picked the number 〈*t_A_*〉 = 6.2 in this interval, to account for about a day earlier exposure that would account for an adjustment for travel, but our results do not change substantially if a slightly smaller value is used. Thus at our chosen value of *f*, this yields

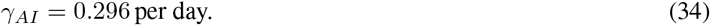

We fix this value in the remainder of the work. Note that all rates will be measured in days.

In a similar manner, we estimate the parameter *γ_IR_*, the transition rate from infected to recovered. The February 2020 joint WHO-China report [25] found an average recovery time of 2 weeks at the time of the onset of symptoms for mild cases, and 3–6 weeks for severe cases. In our model, we do not distinguish between the types of symptomatic individuals, and hence we roughly estimate an average 3 week recovery period. Hence

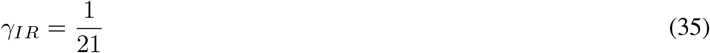

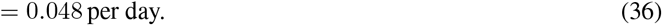

The parameter *f* represents the fraction of SARS-CoV-2 infections that become symptomatic. Current reports suggest that this parameter is highly variable, with analyses on different data sets yielding between 20% – 95% of positive tested cases being symptomatic (so that *f* ∈ [0.2,0.95]) [26, 27, 28, 29, 30, 31, 32, 33, 34, 35]. As of May 2020, the CDC estimates the asymptomatic proportion of positive cases to be 35% [36], so we fix *f* as

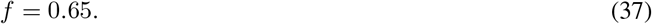

Finally, we can estimate *δ*, the disease mortality rate, using the fraction of reported deaths with respect to the total disease numbers. Using global data reported on the John Hopkins dashboard [37] on 03/29/2020, there were a total of 33876 deaths and 717656 reported cases, so that

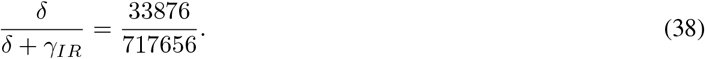

Using the above value of *γ_IR_* in (36), we thus have that

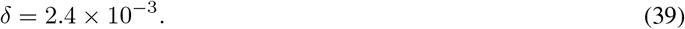

The remaining parameters are *β_A_, β_I_, ϵ_A_*, and *ϵ_S_*. For simplicity, we assume that the effect of social distancing the susceptible, asymptomatic, and symptomatic individuals is symmetric, so that

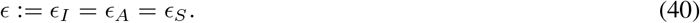

The values of *β_A_* and *ϵ* are calibrated to reported *R*_0_ values. Using data from PolicyLab at CHOP for the Bronx, NY, we use an *R*_0_ = 5.6 when *h*_2_ = 0 (corresponding to 3/23/20), and *R*_0_ = 1.4 when *h*_2_ = 0.5 (corresponding to one week later on 3/30/20) [38]. *Β_I_* is calibrated from *β_A_* using:

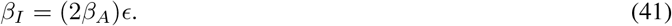

This assumes that symptomatics are twice as infective as asymptomatics [39]. The scaling by *ϵ* indicates that symptomatic individuals are assumed to socially distance.

## 3 Results

### 3.1 Basic Reproduction Number, *R*_0_

A central subject in the analysis of epidemiological models concerns the stability of a “disease free steady state” (abbreviated DFSS from now on), in which all infective populations are set to zero. Stability of the DFSS means that *small perturbations* of the DFSS, that is to say, the introduction of a small number of infectives into the population, results in exponential convergence back to the DFSS. In other words, the infection does not take hold in the population. Mathematically, this means that the linearization at the DFSS is described by a matrix in which all eigenvalues have negative real part (a “Hurwitz matrix”). Conversely, if the DFSS is unstable, then the infection will initially expand exponentially. It is important to realize, however, a very subtle and often misunderstood fact. *Instability of the DFSS does not necessarily imply that the infection will keep increasing forever*. Linearized analysis is only *local*, and says nothing about behavior over long time horizons, because nonlinear effects can dominate once the system is away from the DFSS; indeed, we will show that we find this phenomenon in our model (provided that social distancing directives are introduced).

A fundamental and beautiful mathematical result is that the DFSS is exponentially stable if and only if the *basic reproduction number R*_0_ is less than one. Intuitively, *R*_0_ is the average number of new infections that is caused by a typical individual during the period that this individual is infective. Mathematically, *R*_0_ is defined as the dominant eigenvalue of a certain positive matrix, called the *next generation matrix* [40, 41]. We briefly explain this method in Appendix A, and therein derive that for our six-compartment SIR model in equations (8)-(13):

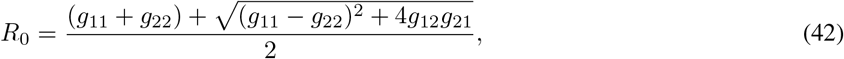

where *g_ij_* represents the (*i, j*) –entry of the next generation matrix *G* evaluated at the DFSS. As we derive in Appendix A:

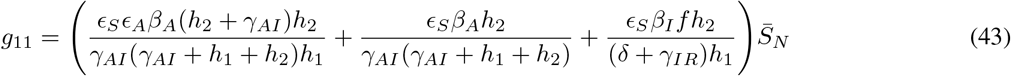

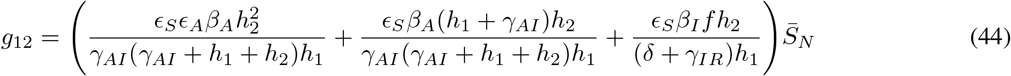

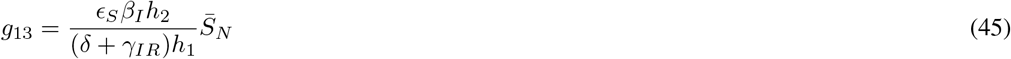

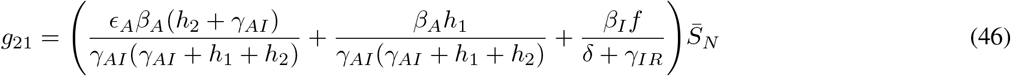

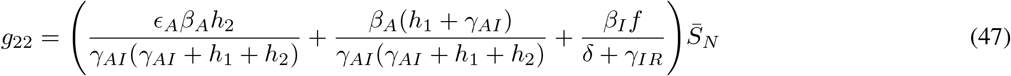

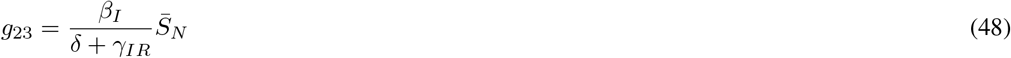

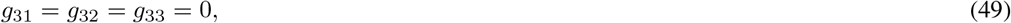

where 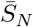 represents the value of *S_N_* at the DFSS.

We begin here by studying the value of *R*_0_ under various scenarios of disease progression. We distinguish these scenarios by a number we call *R*^*^ that gives the percent of the population that has “recovered” from the disease^1^. Herein we assume all recovered have developed immunity, though that assumption could easily be removed. One can interpret *R*^*^ =0 as the very earliest stage of the pandemic, when there are no recovered individuals in the population. On the other extreme, *R*^*^ = 1 indicates every individual has recovered. Because we are assuming all recovereds stay recovered, increasing values of *R*^*^ correspond to increasing values of time.

In Fig. 3, we show how *R*_0_ predicted by our model changes as a function of the fraction recovered *R*^*^ and the rate of social distancing *h*_2_. We observe that at sufficiently early stages of the pandemic (*R*^*^ < 0.3), *R*_0_ > 1 unless social distancing is implemented at a rate faster than *h*_2_ = 0.5, which is already a very rapid rate. (The contact rescaling factor CoRF is kept constant, as explained in the Introduction). While it is easy to interpret this as saying controlling the disease is hopeless unless society could act unrealistically quickly, this is not the case. As discussed above, instability of the DFSS does not characterize global temporal behavior. It does tell us that an “overshoot” and headline-grabbing infection will initially take hold. Thus, social distancing directives will initially appear to have failed in their intended effect. But, as time goes on and individuals limit their contacts and more individuals recover, social distancing can eventually result in *R*_0_ < 1, which would result in the epidemic dying out exponentially.

**Figure 3:**
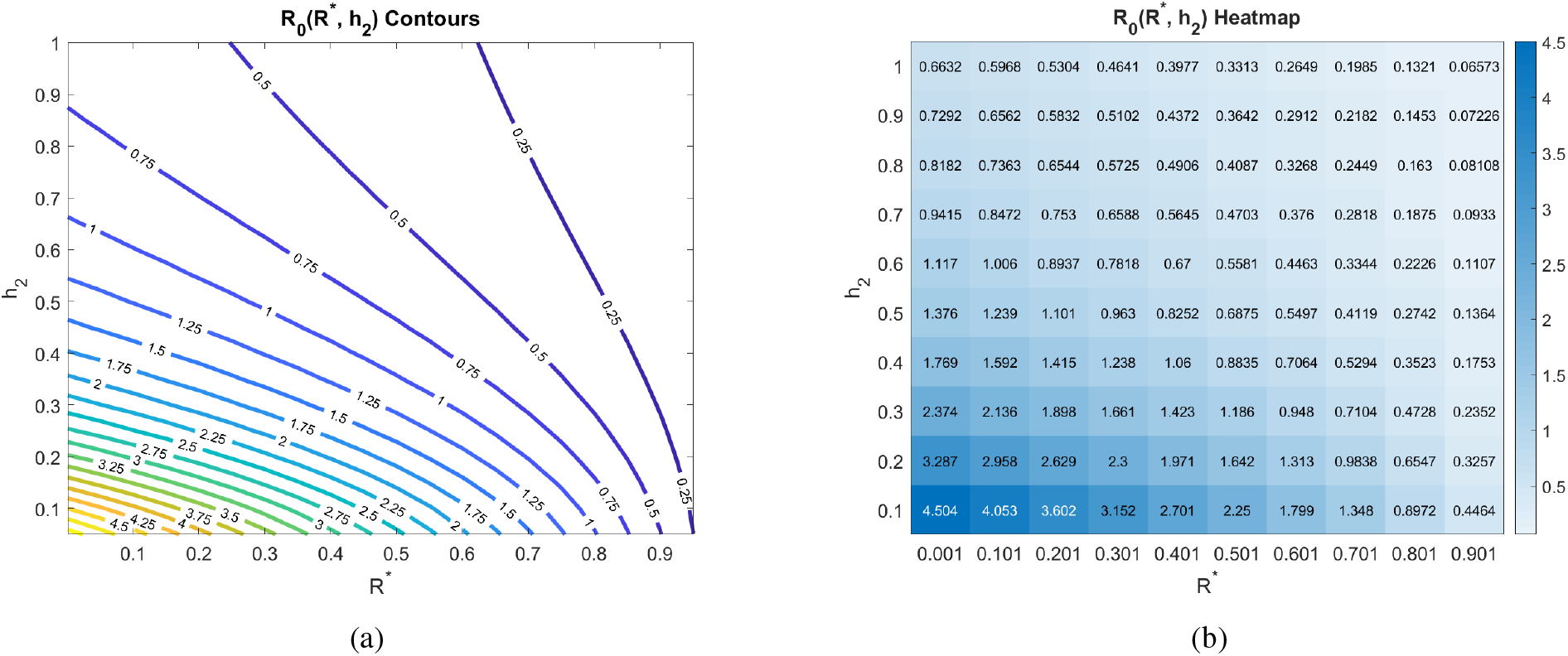
Basic reproduction number as a function of the social distancing rate parameter *h*_2_ and *R*^*^, the fraction of the population that is immune. All other parameters as in Table 2.

Even though it is based on linearization, we found out that analysis based on *R*_0_ is in excellent agreement with simulations, and hence we use formulas for *R*_0_ as a function of *R*^*^ (and of other parameters in the model as well) to understand how sensitive *R*_0_ is to different social distancing rates, the point in time when such directives are introduced (as quantified by *R*^*^), and other parameters. Also note that, even when *R*_0_ > 1, social distancing can still “flatten the curve”, as we show in Section 3.2. This means that the peak infection levels will be lower, which reduces the stress on the healthcare system.

To explore the sensitivity to *h*_2_ further, we note that the limit of *R*_0_ as *h*_2_ → ∞ to can be written as 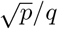, where

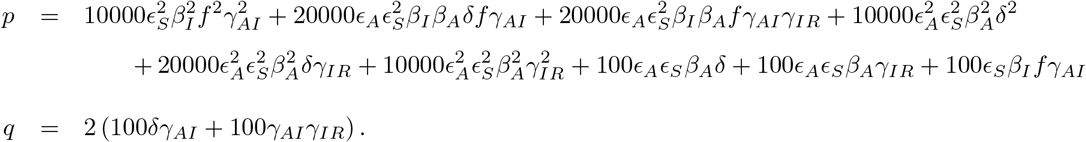

The formulas follow from the explicit calculation of *R*_0_ given in Appendix A. We show in Fig. 4 plots of *R*_0_ as a function of *h*_2_ for *R*^*^ =0 (on the interval *h*_2_ ∈ [0,1]), as well as its derivative and its differential sensitivity, defined intuitively as “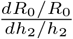” and formally as 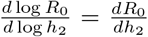. The fact that this sensitivity rapidly approaches zero means that after a threshold rate of social distancing, *small relative changes in the rate of social distancing have essentially no effect on relative changes in R*_0_.

**Figure 4:**
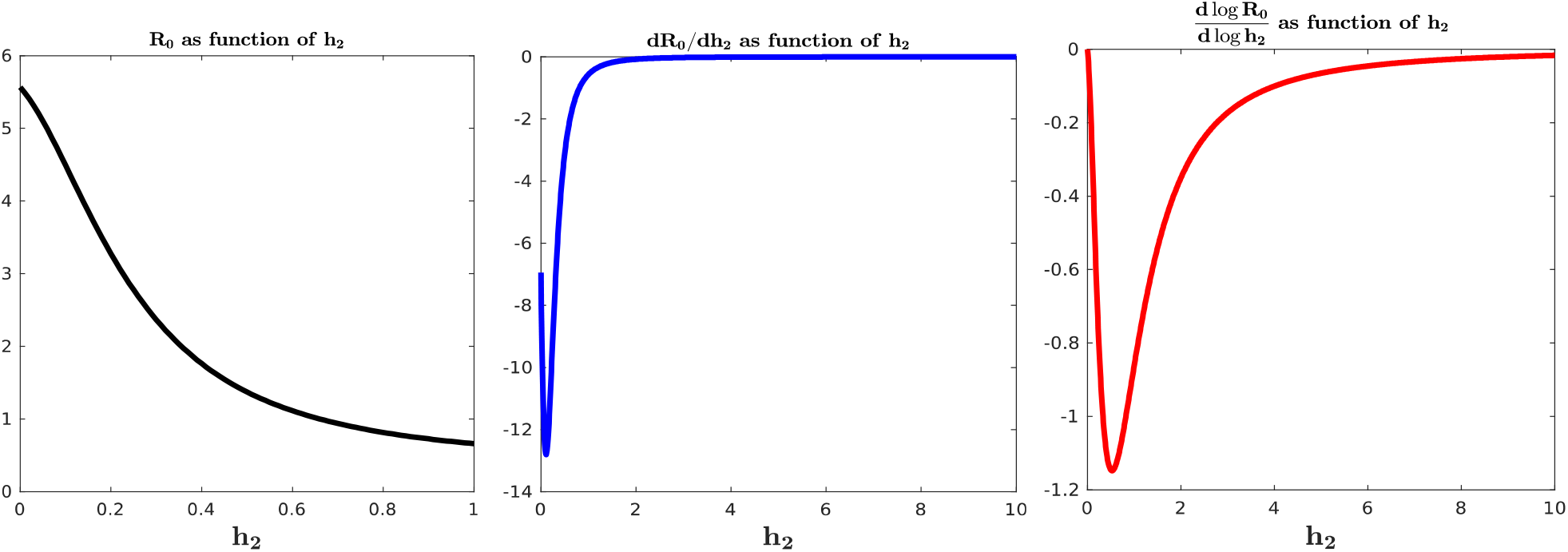
Plots of *R*_0_, derivative d*R*_0_/*dh*_2_, and differential sensitivity 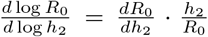, all as functions of *h*_2_ at *R*^*^ = 0. Different ranges picked for clarity. *dR*_0_/*dh*_2_ and differential sensitivity converge to zero.

Mathematically, it is interesting that the derivative of *R*_0_ (and also the sensitivity) has a local minumum, in other words 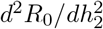 can change sign. This is necessary because 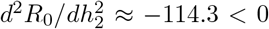 at *h*_2_ = 0, but this second derivative cannot stay negative since *R*_0_ is bounded below. Taken together, these plots further confirm that increasing the rate of social distancing beyond a certain threshold does not result in significant changes to *R*_0_.

We proceed by exploring the sensitivity of *R*_0_ to various combinations of parameters, all including the social distancing rate *h*_2_. The first parameter we consider is *f*, which determines the fraction of asymptomatic individuals that progress to having disease symptoms. Varying this parameter is important, as different studies have reached different conclusions about its value. As Fig. 5 indicates, the value of *R*_0_ is sensitive to the fraction of individuals that develop symptoms, whether we are at an earlier (*R*^*^ = 0) or later (*R*^*^ = 0.25) stage of the pandemic. In particular, a larger likelihood of transitioning from asymptomatic to symptomatic increases *R*_0_. This occurs because both asymptomatics and symptomatics spread the disease, so spending time in both A and I means an individual has more time to spread the disease than if an individual transitions directly from the asymptomatic pool to the recovered pool. If instead of using the CDC estimate that *f* ≈ 0.65 [36], we used earlier data from the Diamond Princess cruise ship [26] of *f* ≈ 0.821, without any social distancing directives, this increase in *f* would increase *R*_0_ from 5.6 to 6.3 at our baseline parameters. Yet another data set out of Italy suggested *f* may be closer to 0.57 [35]. Without any social distancing directives, this reduction in *f* would reduce *R*_0_ from 5.6 to 5.3 at our baseline parameters. While these changes are not drastic, we do note that *R*_0_ is more sensitive to *f* as *h*_2_ increases, as can be seen in Fig. 5 by observing the increasing steepness of the contours as *h*_2_ increases. Further, the possibility that there may be different strains of SARS-CoV-2 could mean that *f* varies depending on the dominant strain in a region [42].

**Figure 5:**
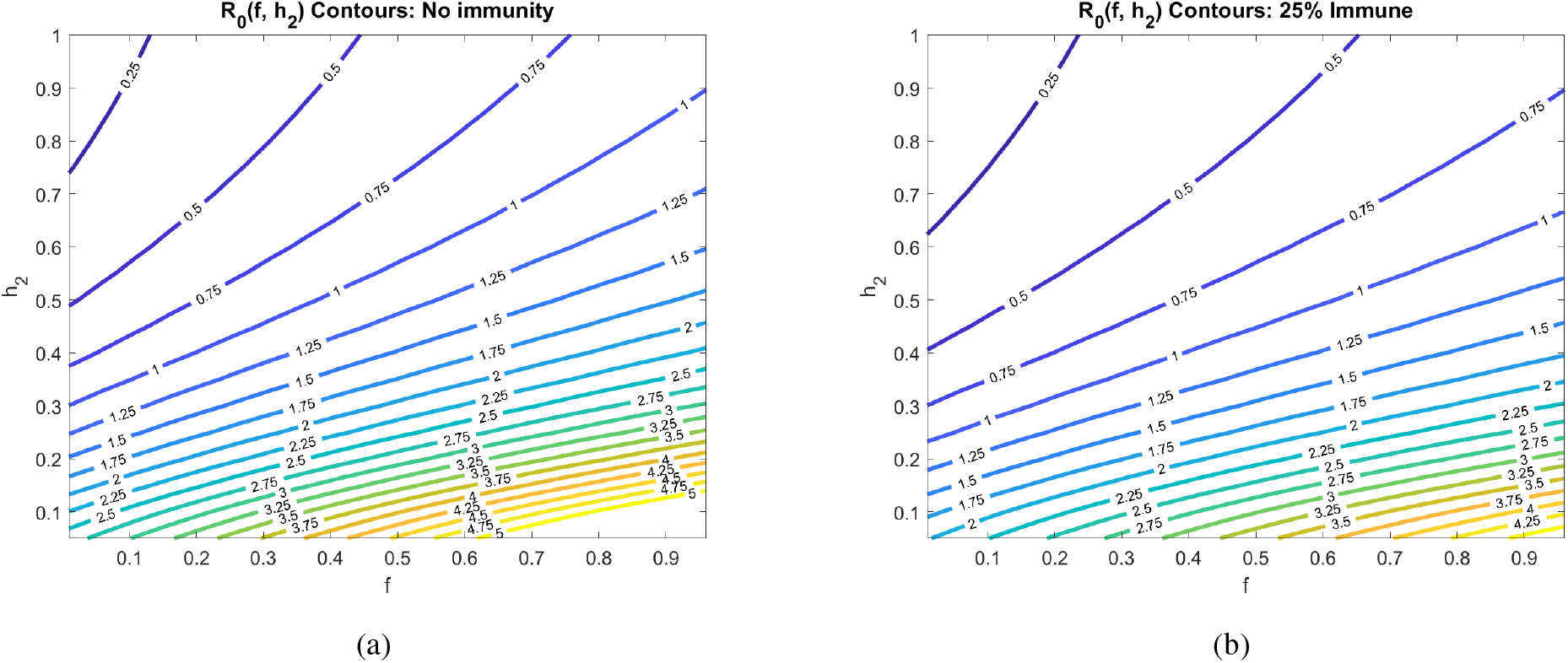
Basic reproduction number as a function of the social distancing rate parameter *h*_2_ and fraction of individuals who become symptomatic (*f*) at different pandemic stages. All other parameters as in Table 2.

Another uncertainty surrounding COVID-19 is how infectious the asymptomatic individuals are, which we call *β_A_* in our model. In Fig. 6, we explore how the contagion level of the asymptomatics influences *R*_0_ under varying rates of social distancing. We observe that if asymptomatics are not very contagious (*β_A_* is small), then *R*_0_ is not very sensitive to social distancing directives as measured by *h*_2_ (*R*_0_-clines are almost vertical). This can be explained by our assumption that symptomatic individuals are assumed to socially distance themselves, and therefore have minimal interaction in our model with susceptibles. When this is the case, the disease is mainly spread by non-socially distanced asymptomatics. And, if the transmission rate from these individuals is small, socially distancing the asymptomatics and susceptibles has little impact on the progression of the disease. If, on the other hand, *β_A_* is sufficiently large, then asymptomatics can fairly readily spread the disease, and we see a much stronger impact of social distancing on *R*_0_ (*R*_0_-clines get much less steep as *β_A_* increases).

**Figure 6:**
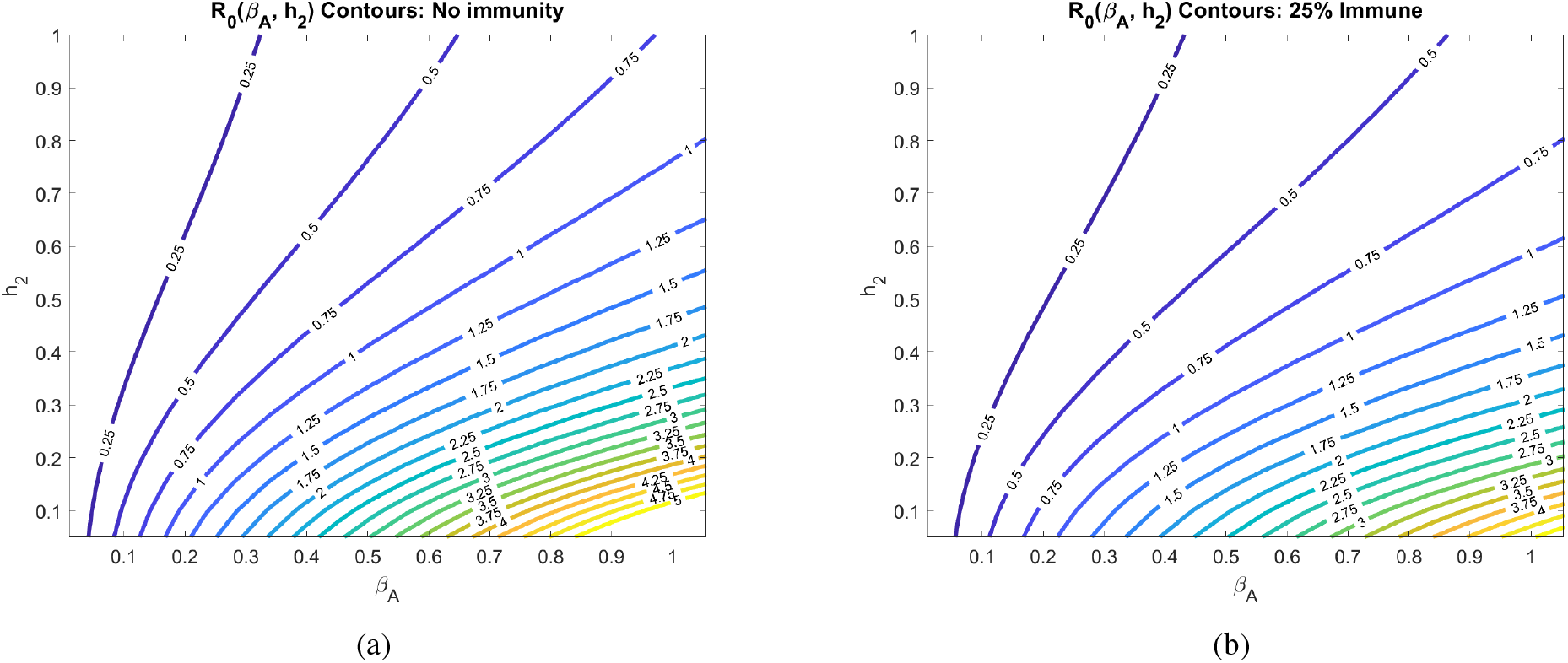
Basic reproduction number as a function of the social distancing rate parameter *h*_2_ and infectivity rate of asymptomatics *β_A_* at different pandemic stages. All other parameters as in Table 2.

Another major assumption of our model is that social distancing reduces the transmission rate of the disease by a factor called the contact rescaling factor (CoRF). We formulate our model so that socially distancing the susceptibles and the asymptomatics (note, infectives are assumed to be socially distanced) are described by different CoRF values of *ϵ_S_* and *ϵ_A_*, respectively. However, in all calculations and simulations, we assume that the extent that social distancing reduces the transmission rate is the same independent of whether an individual is susceptible, asymptomatic, or infected. That is, we take *ϵ* := *ϵ_S_* = *ϵ_A_*. The value of the CoRF *ϵ* is another way to measure social-distancing directives. While *h*_2_ describes rate of social distancing, *ϵ* describes the severity of the measures. While not realistic for a disease like COVID-19, if socially distancing meant an individual was exposed to nobody else, the contact rescaling factor *ϵ* would be 0. Intuitively, and as we quantitatively demonstrate in Fig. 7, at very small *ϵ*, the rate of social-distancing *h*_2_ is less important. Social-distancing is still needed in order for *R*_0_ to be less than 1, as it reduces the transmission from the distanced symptomatic individuals to susceptibles. In particular, *R*_0_ drops below 1 at the rate of *h*_2_ = 0.3 when there are no recovered in the population, and at *h*_2_ = 0.24 when 25% of the population has recovered.

**Figure 7:**
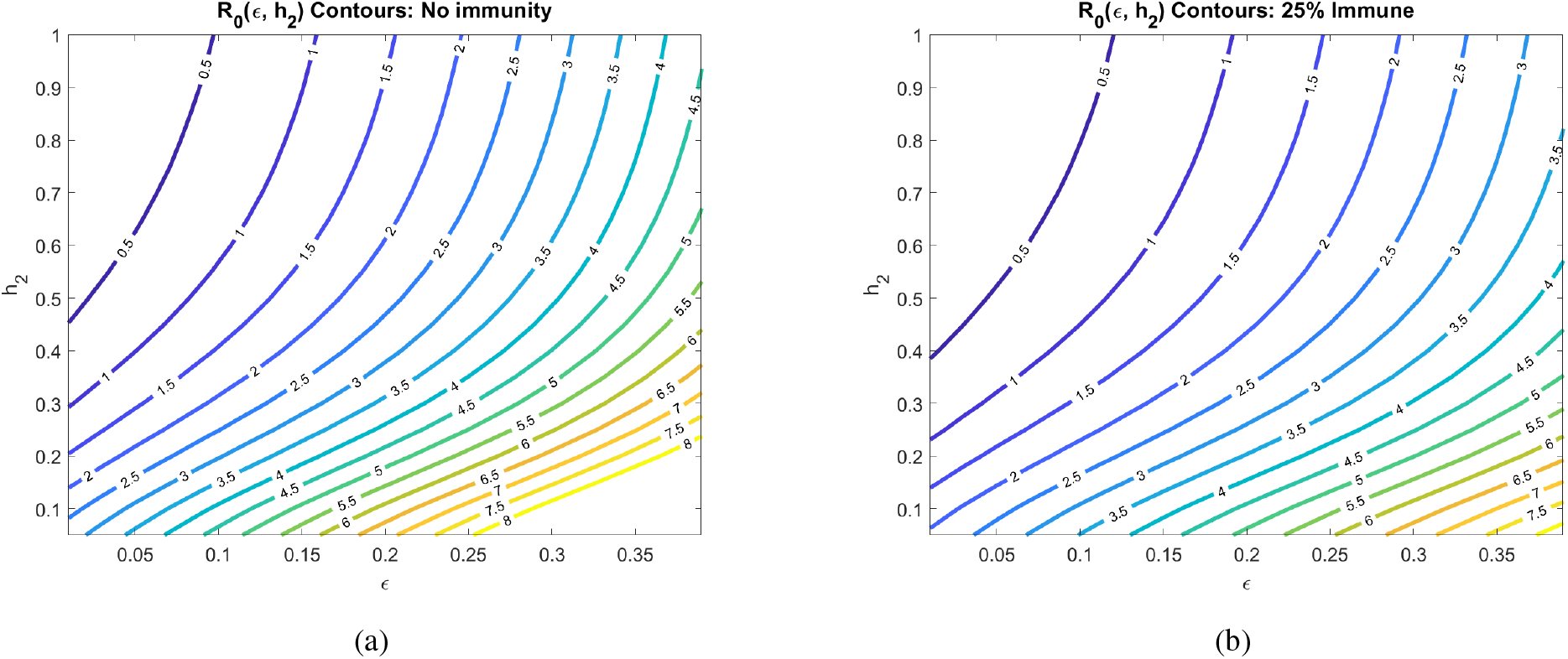
Basic reproduction number as a function of the social rate distancing parameter *h*_2_ and the contact rescaling factor (CoRF) *ϵ* at different pandemic stages. CoRF measures the impact of social distancing on infectivity rate. All other parameters as in Table 2.

Increasing the CoRF *ϵ* can be thought of as increasing the number of contacts socially-distanced individuals have. With all other parameters fixed as specified in Table 2, we see that the social distance rate *h*_2_ can only result in an *R*_0_ < 1 at the early stages of the pandemic (for *h*_2_ ≤ 1) if the CoRF *ϵ* is less than 0.15. Beyond this value, even socially-distanced individuals have too many contacts, and *R*_0_ > 1. Intuitively, we see that *R*_0_ quickly increases as the CoRF increases. *This shows that, even if individuals act very quick to socially distance (there is a large h*_2_*), if they have too many contacts while social distancing, social distancing will not be sufficient to drive R*_0_ < 1.

**Table 2:**
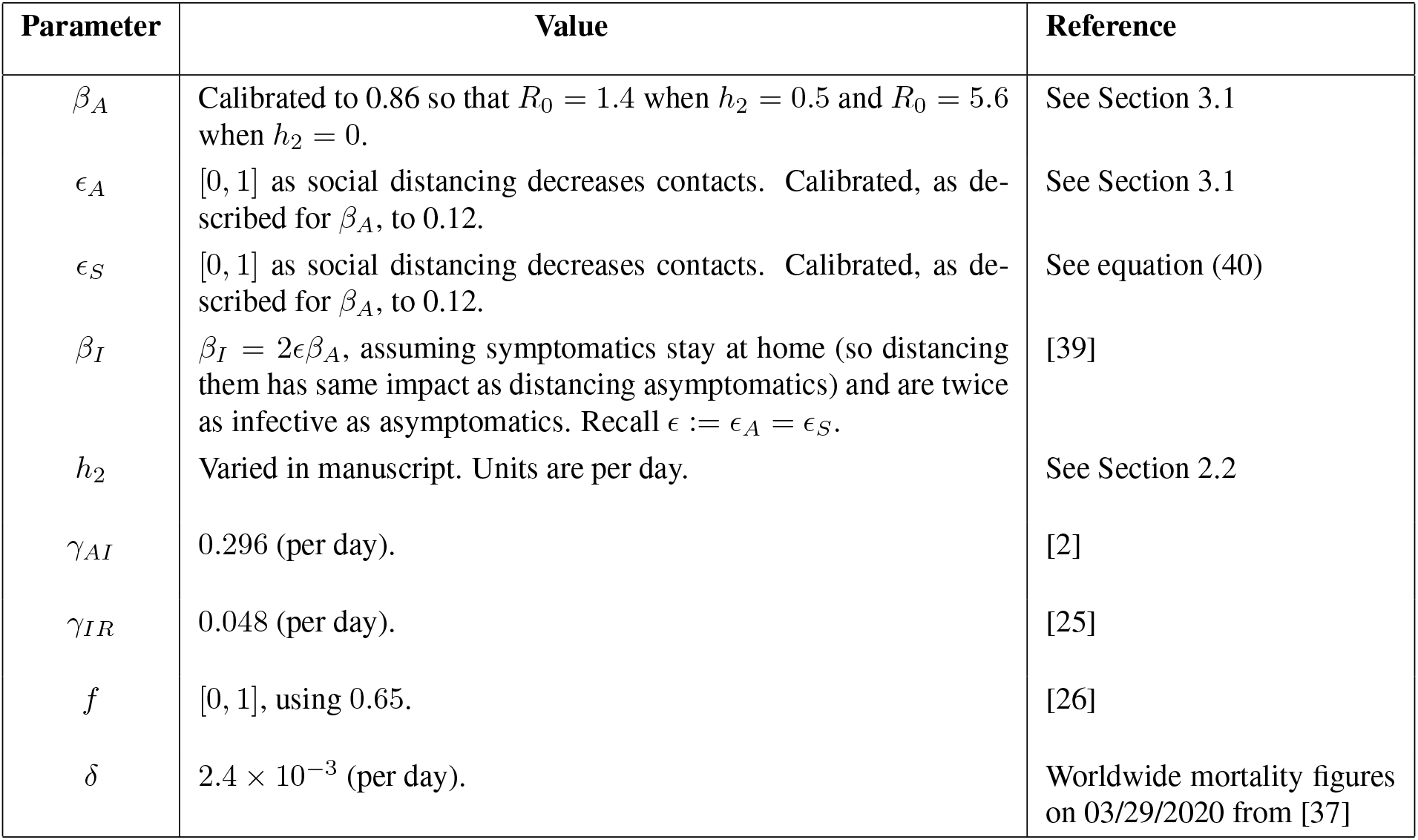
A list of estimated parameters values to be used in our simulations.

### 3.2 Time-varying social distancing (*h*_2_(*t*))

We now investigate how dynamic social distancing protocols affect the spread of the epidemic in the six equation SIR model introduced in Section 2.1.

#### 3.2.1 No distancing dynamics

We begin with the predicted outbreak dynamics in the case of no implemented social distancing. As discussed in Section 3.1, with parameters estimated from data, we expect

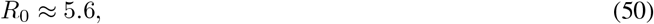

so that we expect the disease to spread rapidly throughout the population. No social distancing implies that

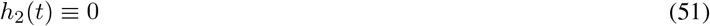

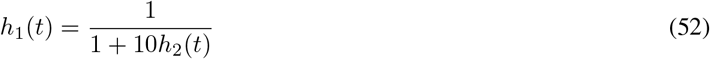

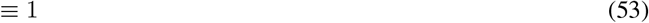

Initial conditions consist of all susceptible individuals socially non-distanced, no population immunity, and a small number of symptomatic individuals (100 in 10 million). More precisely,

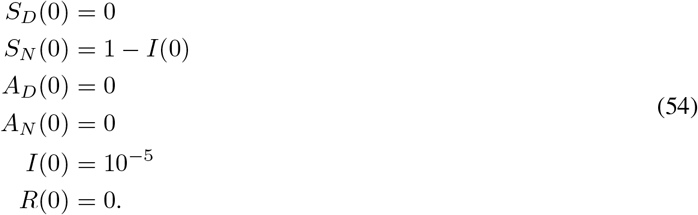

Of course, in reality it is likely that a number of asymptomatic people also exist in the population at this time (*t* = 0), but for simplicity we ignore them. Note that *I*(0) was taken to be consistent (as an order of magnitude) with the reported cases in New York City (NYC) before a state of emergency was declared (89 cases on March 7, with a total NYC population of approximately 8.4 million). These will be the initial conditions used throughout the remainder of the manuscript, as we compare intervention strategies based on responses to the above infection data at day *t* = 0.

The results of simulating the model in equations (8)- (13) for 180 days are provided in Fig. 8. This will serve as a baseline for the severity of the outbreak in a “worst-case scenario.” Social distancing strategies will be compared to these worst-case figures, some of which we highlight below:

1. The symptomatic infected population (*I*) reaches a peak value of 40%.
2. The time to this symptomatic peak is approximately 30 days.
3. The model predicts that a peak of 60% of the total infected population (symptomatic and asymptomatic) will occur at day 24. As a reminder, these numbers are based on parameters calibrated to the outbreak in the Bronx NY, a locale hit very hard by the virus.
4. By day 70, 90% of the original the population will be recovered (assuming recovered individuals do not lose immunity).
5. Over 3% of the population will die during the outbreak (3.11%).
6. Compartments *S_D_* and *A_D_* never comprise any percentage of the population. This is because of the initial conditions, and because no distancing guidelines have been issued in this “worst-case scenario”.

**Figure 8:**
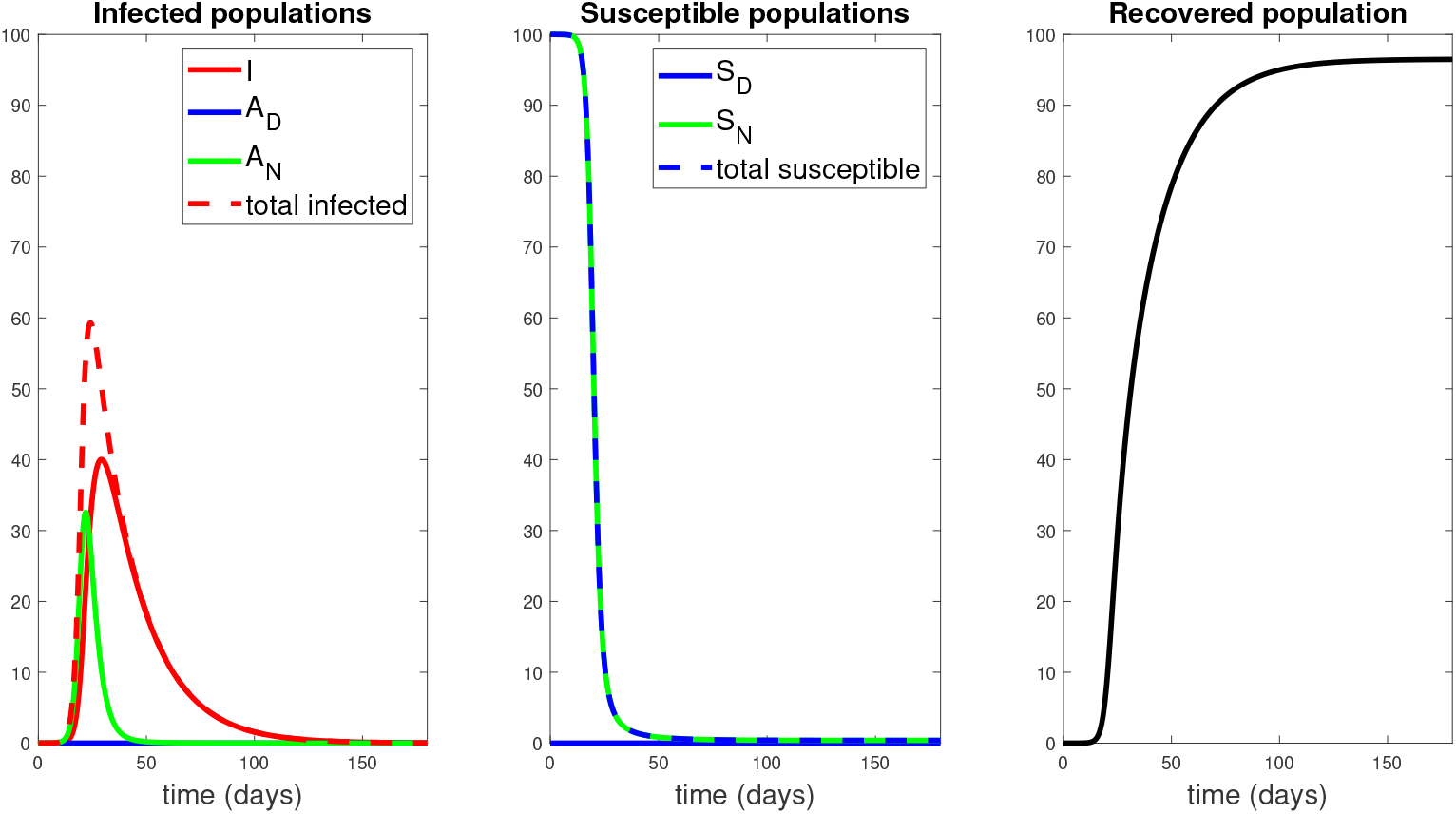
Population responses in the absence of treatment. Here *h*_2_(*t*) ≡ 0, i.e. no social distancing has been implemented. We simulated the model introduced in Section 2.1 for 180 days. Note that the infected symptomatic population (*I*, solid red curve) comprises about 40% of the population by day 30. Units of vertical axes are percentage of initial population.

#### 3.2.2 Delayed response

We first investigate disease dynamics in response to delayed social distancing protocols. We assume that *h*_2_(*t*) takes the following form:

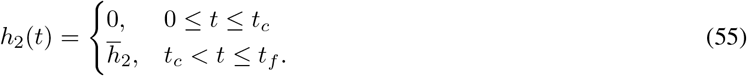

That is, we assume that social distancing does not occur until an implementation time *t_c_*, after which *h*_2_(*t*) is kept at a constant value 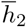. For a visualization, see Fig. 9.

**Figure 9:**
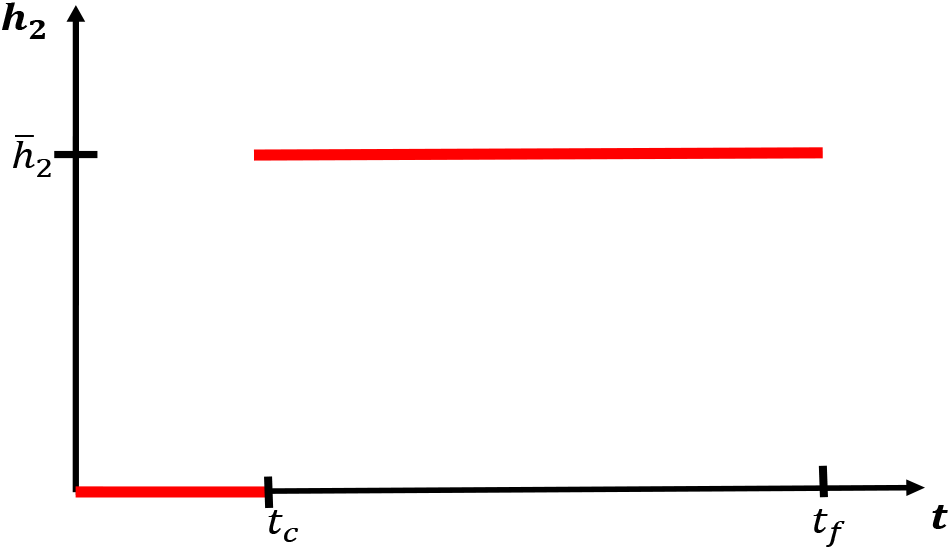
Social distancing control as a function of the delay *t_c_*. Functional form for *h*_2_ used in Figs. 10, 11. See also equation (55).

Since many policies are implemented in a 48 hour (2 day) window, we set

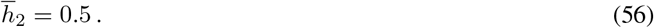

Fixing all other parameters as in Table 2, we simulate the model for a range of policy activation days t_c_; results are shown in Fig. 10. Here we observe an apparent “flattening of the curve,” *if the distancing was enacted quickly enough*. That is, if policies were enacted too late, social distancing has little effect on the course of the outbreak. This can be observed by noticing that the response to a delay of 25 days is nearly identical to one with no distancing imposed. This is hardly surprising, since if a society waits too long to start socially distancing, the disease will have already spread through much of the population. However, if the delay is short enough (i.e. the response quick enough), we see a significant reduction in the peak of the infected population (3% at the peak for a *t_c_* = 5 days, compared to the worst-case of 40%). Furthermore, there seems to be a critical “window of opportunity” for commencing social distancing: the difference between waiting 15 and 20 days is quite striking (peak of 7% in the former, up to 30% in the latter). *Notice that this window of opportunity ends at a time (about* 15 *days) that is much earlier than the time at which infections would have peaked in the absence of control measures (about* 30 *days). We think of (roughly)* 15 *days as a* critical implementation delay (CID).

**Figure 10:**
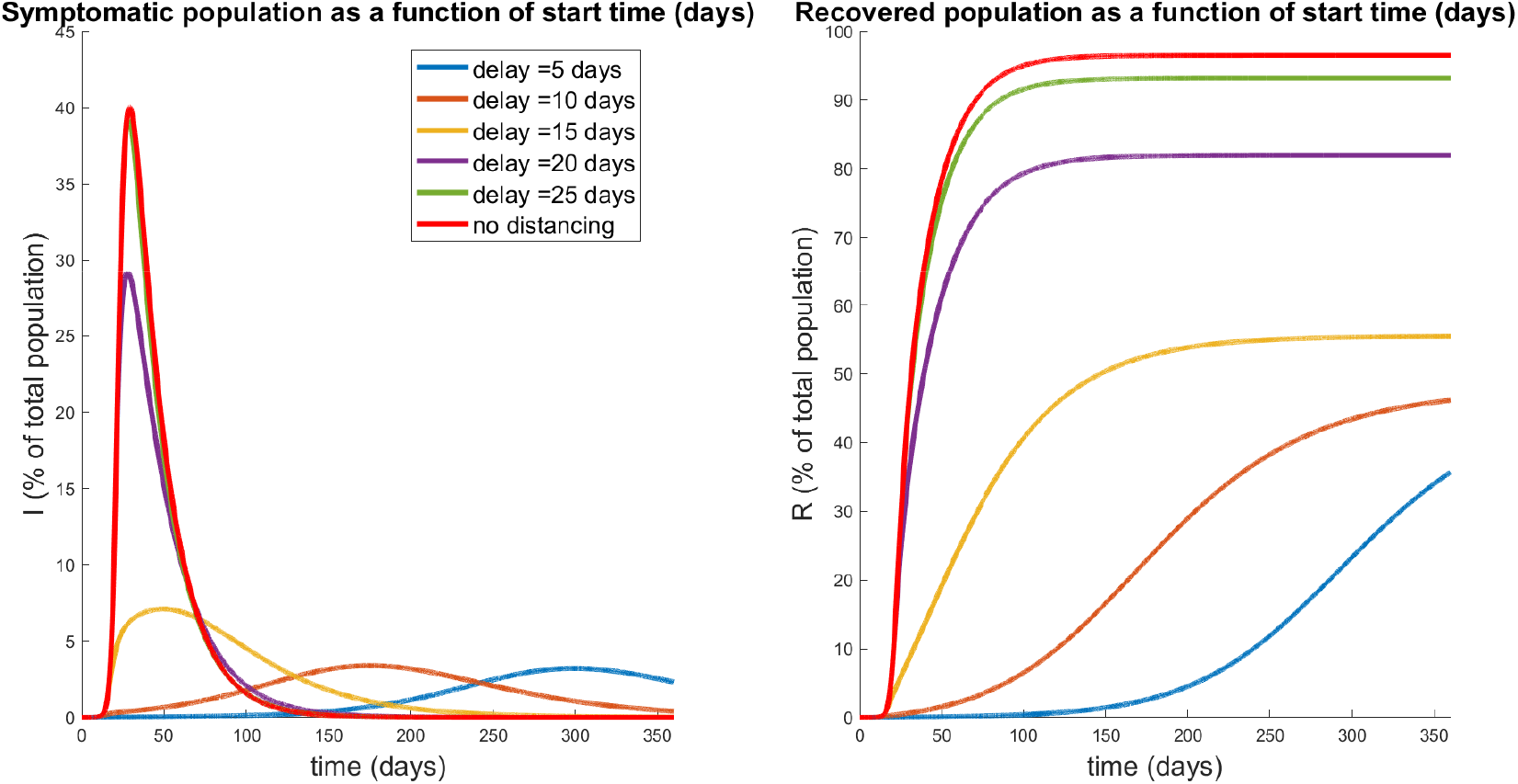
Population response for a treatment window of 360 days with varying start time *t_c_* of social-distancing protocol (see Fig. 9). Left panel denotes the symptomatic (*I*) temporal response; right indicates the recovered percentages for each policy. Red curves correspond to no social distancing (see Fig. 8). Note that a delay of 25 days is hardly discernible from no social distancing, while a significant response transition occurs for delays shorter than 15 days.

We investigate this further by plotting both the peak symptomatic population and the time to this peak in Fig. 11, where social distancing is begun at *t* = *t_c_* days, for *t_c_* = 0,1,2,…, 35. This provides a quantification of what we saw in Fig. 10: if the delay is relatively small or large, the response (measured as peak infected percentage) is robust to the delay. However, there is a critical window about which a “bifurcation” occurs. For our parameters, the bifurcation value appears to be approximately 2 weeks. In those first two weeks, if social distancing is begun, the outbreak will be sharply inhibited. However, near this critical value, delaying even a few extra days could drastically increase the total number of symptomatic individuals. For example, waiting 17 days yields a peak of 14% symptomatic, while waiting an extra week increases the peak to over 35%. Thus we see that policies will be effective in a certain window, and that it is critical to implement them within that window. Indeed, delaying even by a few days *outside of that window* could severely increase the total number of infections.

**Figure 11:**
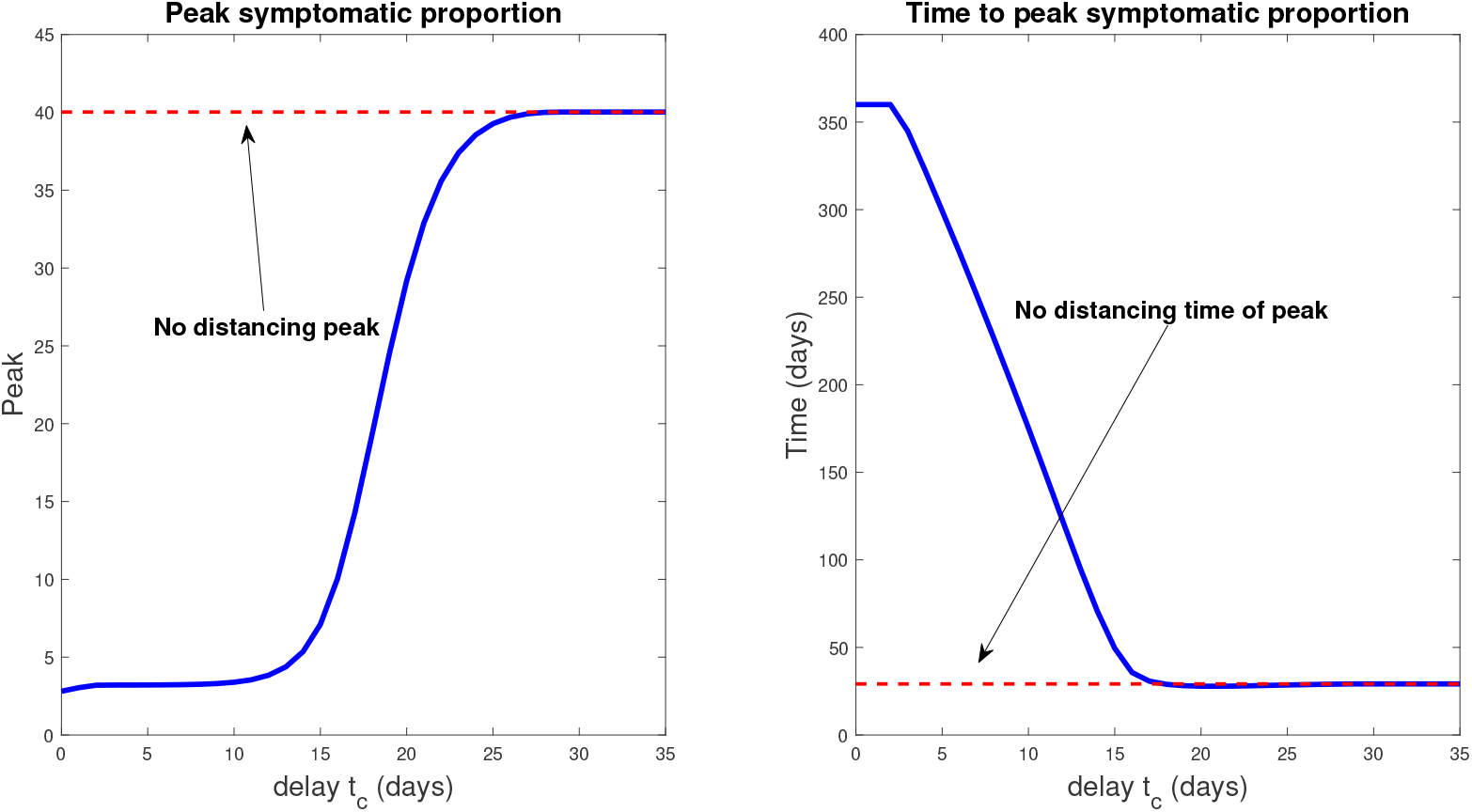
Peak infected population percentage (left) and time to this peak (right) when social distancing is delayed. Here the horizontal axis represents the delay (from time *t* = 0), i.e. the value *t_c_* in equation (55). The dotted red line denotes the corresponding values when no social distancing is enacted (Section 3.2.1). Note the rapid increase in peak symptomatic population beginning around 15 days, which we term a *critical implementation delay (CID)*.

We also note that the *time to peak number* of symptomatic individuals (right panel, Fig. 11) increases the sooner the distancing procedure is implemented. This is intuitive, and combined with the previous result says that the more quickly social distancing is enacted, the longer you will have to deal with a smaller number of sick individuals. However, the number of sick individuals is relatively constant *up until a certain time delay*, where once passed, there will be many more (over ten times) the number of sick people in the population at its worst moment. Hence, on the policy level, if the existence of the infection is known sufficiently early, it is okay to take some time to plan a strategy, but once decided, it must be implemented quickly and efficiently. On the other hand, *implementing even faster than the CID time results in the peak infections being postponed by a huge margin*, thus giving more time for the development of vaccines and treatments. For example, a delay of 15 days has a peak infection early on in the epidemic (at about 50 days), while implementing the same distancing policy 10 days earlier delays the peak until approximately 1 year. In both cases, the peak infected populations are similar (7% compared to 3%, still both substantially smaller than the predicted 40% non-distanced dynamics), but the time scales over which the peak occurred are much different. Thus, one must take into account both factors (peak, and time-to-peak) when designing distancing strategies.

#### 3.2.3 Periodic relaxation

We next investigate the effects of periodically relaxing social distancing protocols. Consider a protocol where social distancing measures are implemented for a fixed time window Δ*t*_on_ followed by a relaxation Δ*t*_off_. The above is then repeated until a final time *t_f_* is reached. We envision a situation where the population is allowed to interact normally for (say) one week, but must then isolate for the following week. Such policies may lessen the economic and psychological impact of extended complete isolation by allowing limited windows in which individuals may work, socialize, etc. For a visualization of a simplified version of such a policy, see Fig. 12. Note that we consider total relaxation (*h_2_* = 0) during Δ*t*_off_, but of course this could be adjusted; here we consider the simplest possible periodic (i.e. metronomic) policy.

**Figure 12:**
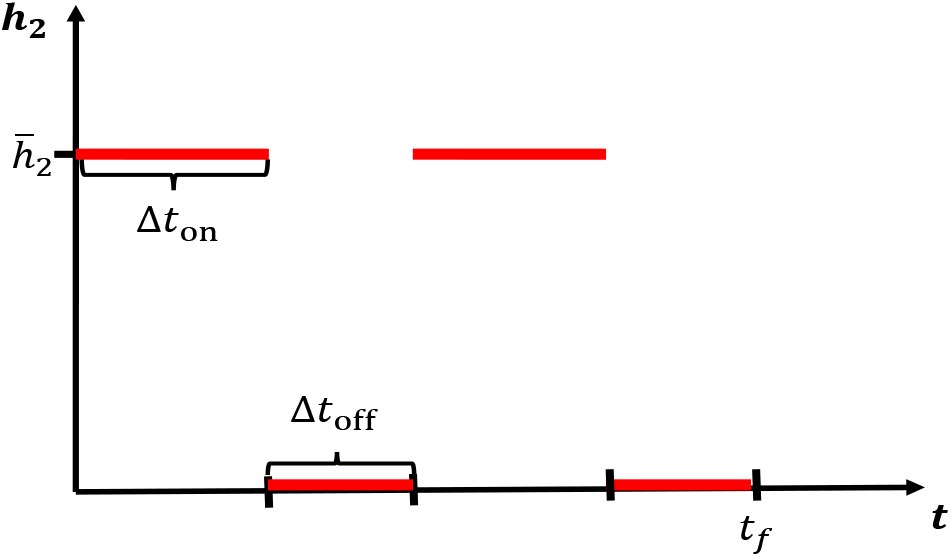
Pulsing of socially distancing protocol. Social distancing is enacted for a time length of Δ*t*_on_ days, followed by a full relaxation for Δ*t*_off_ days. This schedule is then repeated until a time window *t_f_* has been reached.

In Fig. 13, we investigate the dynamical response to several schedules with varying number of weeks of distancing. We assume that the lengths of activation and relaxation are equal, so that

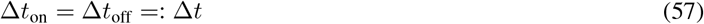

**Figure 13:**
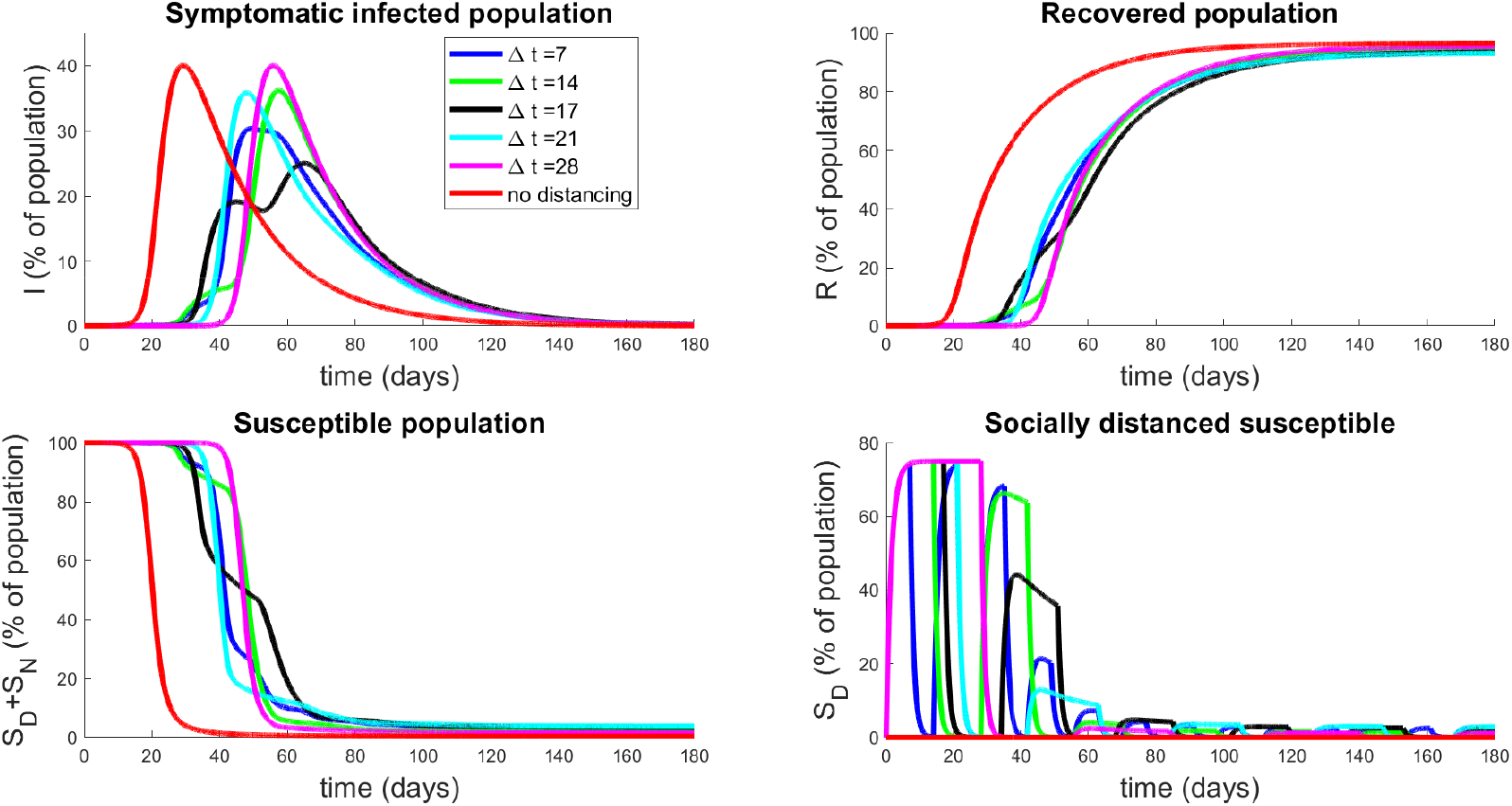
Population response to strategies based on periodic relaxation of social distancing; see Fig. 12. Top left panel denotes temporal dynamics of symptomatic population (*I*) for pulsing strategies with Δ*t*_on_ = Δ*t*_off_ = Δ*t*. Non-socially distanced dynamics (red curves) are provided for comparison. Top right panel is recovered population in time, and bottom panels are total susceptible individuals (left) and socially-distanced susceptible individuals (right). Initial conditions are described in Section 3.2.1.

Note that this restriction allows a relatively unbiased comparison between distancing protocols, since all will have distancing enacted for the same total amount of time. There is a slight discrepancy based on *t_f_*, since the schedules may end at different points of their respective cycles, but this effect is minimal. Hence we conclude that each schedule will have approximately the same economic impact, and hence in the following we only examine the disease response. Note that we fix 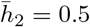 as in Section 3.2.2.

The results presented in Fig. 13 are quite surprising and non-intuitive. Note that they all have delayed the onset of the peak of the epidemic by a similar length of time (all around 45 – 65 days, whereas the epidemic would have originally peaked at around 30 days). However, the degree to which the peak has been suppressed is different among the relaxation schedules. It appears that high frequency pulsing (small Δ*t*) does better than some extended strategies (compare Δ*t* = 7 to Δ*t* = 14,21, 28), but worse than others (compare Δ*t* = 7 to Δ*t* = 17). This seems to indicate that there is some optimal pulsing period. Furthermore, the curve for Δ*t* = 17 days is quite interesting; we see a significant reduction in peak population infection (22%, compared to 30% for Δ*t* = 7 days, and approximately 40% for no social distancing) together with an extended “flattening of the curve”, which does not appear in the others. We also note that all strategies end with similar recovery rates (all above 93%, top right panel, Fig. 13).

To understand the behavior near Δ*t* = 17, we simulate a series of strategies with Δ*t* near this value, and observe how the symptomatic response varies. Results are provided in Fig. 14 for Δ*t* = 16,17,17.6,18,19 days. Note that the peak infected proportion appears to interact with a concavity change near Δ*t* = 17.6, and this interaction causes a significant decrease in the peak together with an extended “flattening” period. However, it is relatively sensitive to the timing, so that a slight error in timing (or a slight variation in parameters) will cause a large increase in peak infected numbers. Another interesting property is apparent in the curve with Δ*t* = 17: we see an initial flattening and even reduction of infections, followed by an increase in the number of symptomatic individuals. Hence for some strategies, the progression may yet worsen even after an apparent downward trend. *Phenomenologically, we see a “bifurcation” between two “unimodal” behaviors in time (earlier* vs. *later peak) that happens through a “bimodal” (two maximal) time behavior (centered around a period of approximately* 35 *days corresponding to* 17.5 *days of distancing and* 17.5 *days of non-distancing)*.

**Figure 14:**
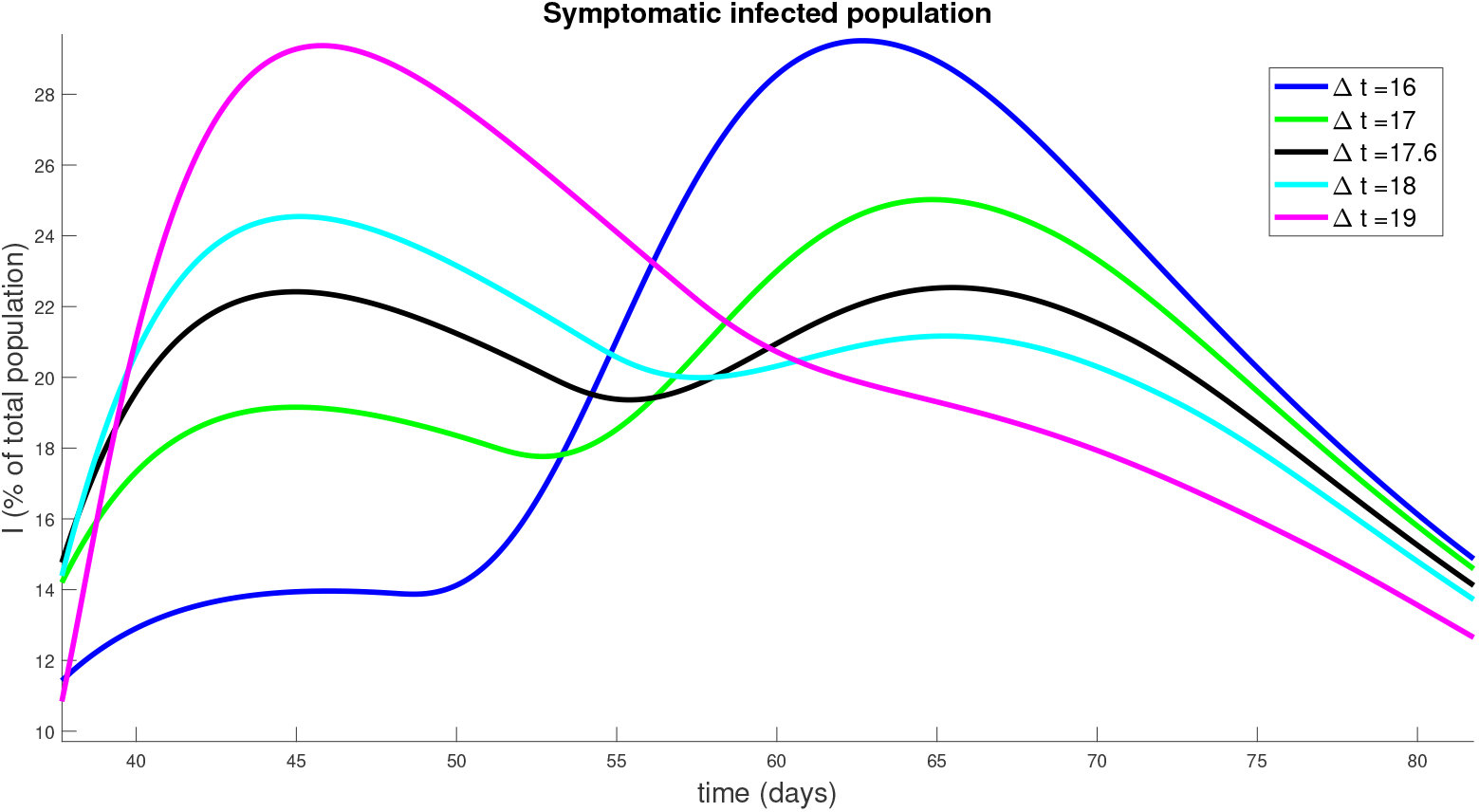
Symptomatic population response to strategies based on periodic relaxation of social distancing (similar to Fig. 13), but for policies with Δ*t* near 17 days.

We also globally investigate the response to different pulsing frequencies (different Δ*t*) on the critical infection measures, namely peak symptomatic individuals and the corresponding time of this peak. A simulation of pulsing strategies with periods ranging from Δ*t* = 1 day to Δ*t* = 70 days is presented in Fig. 15. The left panel denotes a clearly non-monotone global response to different periodic relaxation schedules. Furthermore, we observe a global minimum near Δ*t* = 17.6 days, as discussed previously. Note also the sensitivity to the period: Δ*t* = 17.6 days yields a peak of only 22%, while a slightly longer relaxation schedule of Δ*t* = 20 days produces a peak of over 33%. Hence, designing such strategies is inherently risky, and should be done only when parameter values are precisely known.

**Figure 15:**
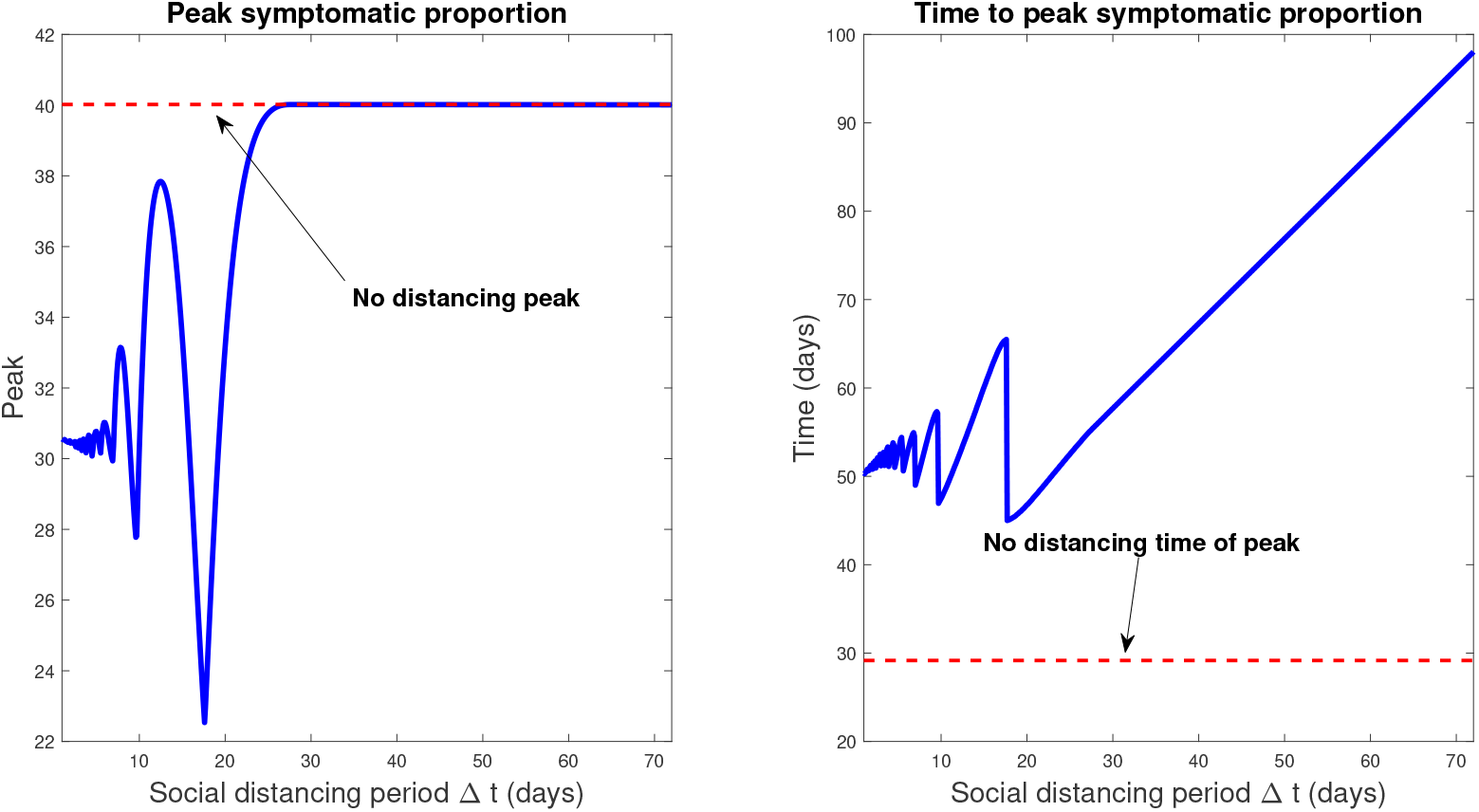
Response of infection dynamics to periodic relaxation for a range of frequencies. Policy period is assumed for 360 days. Left panel denotes peak symptomatic population (percentage) at any one time. Right panel is the corresponding time (in days) when this peak occurs. Initial conditions are described in Section 3.2.1.

#### 3.2.4 Relaxing social distancing

We next investigate the rate at which social distancing policies are eased after a fixed period of time. Such control strategies may be important to prevent a second wave of infection arising soon after policies are relaxed. In this section, we model the effects of a controlled relaxation on outbreak dynamics. The control we consider takes the form of a linear decrease in regulations after a fixed isolation period (*t*_1_ days). The rate of decrease is determined by an end time *t_e_*, after which social distancing is no longer encouraged. Thus, a larger value of *t_e_* corresponds to a slower easing of restrictions. For a visualization, see Fig. 16. We fix

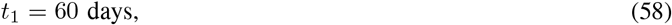

to capture conditions as of May 2020.

**Figure 16:**
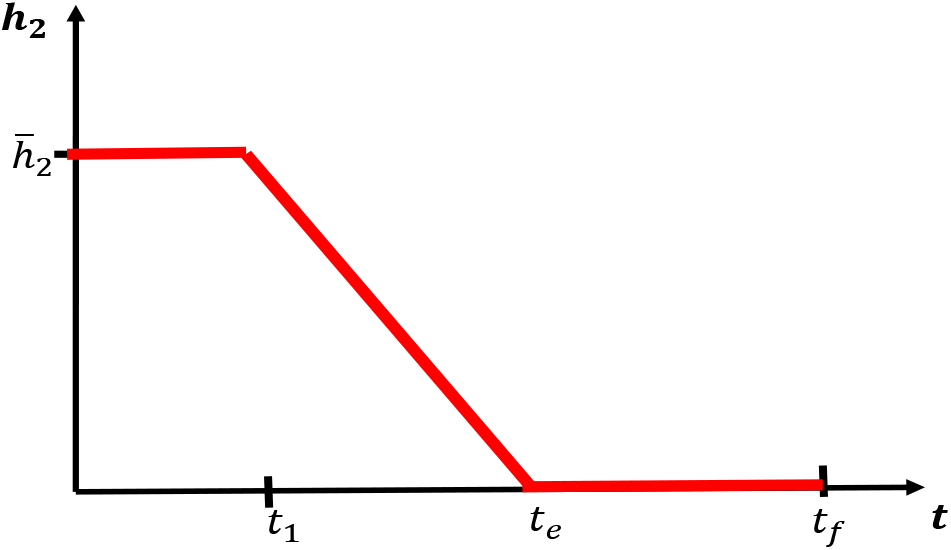
Relaxing social distancing measures after an initial period of strict regulations for *t*_1_ days. Rate is decreased linearly from 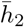 at day *t*_1_ to 0 at day *t_e_*. After day *t_e_*, no distancing regulations are in place.

Results of simulation data for select relaxation rates appear in Fig. 17, while a global characterization is provided by Fig. 18. Note that *t_e_* = 60 days corresponds to immediately turning social distancing off (a step, i.e. infinite slope), while *t_e_* = 360 corresponds to a relaxation rate of slope 0.0017. Results indicate that gradual relaxation does have a significant effect on “flattening the curve” in that it results in a lower peak infected population over a larger time interval (compare *t_e_* = 60 to *t_e_* = 360 in Fig. 17). Fig. 18 further supports this claim, as we compute little variation in the peak for small relaxation times (corresponding to more quickly ending protocols), but that a substantial decrease in symptomatic burden is obtained as *t_e_* is increased. Indeed, for a gradual relaxation over a one year period (360 days), we see a peak symptomatic population of only 19%, which is down significantly from immediate re-openings after 60 days (40%). Note that the latter schedule does not result in any significant peak mitigation when compared to the policy of no social distancing: it merely delays the same peak by approximately 56 days. Hence it seems crucially important to gradually relax social distancing guidelines, as gently as is economically feasible, to help mitigate the outbreak.

**Figure 17:**
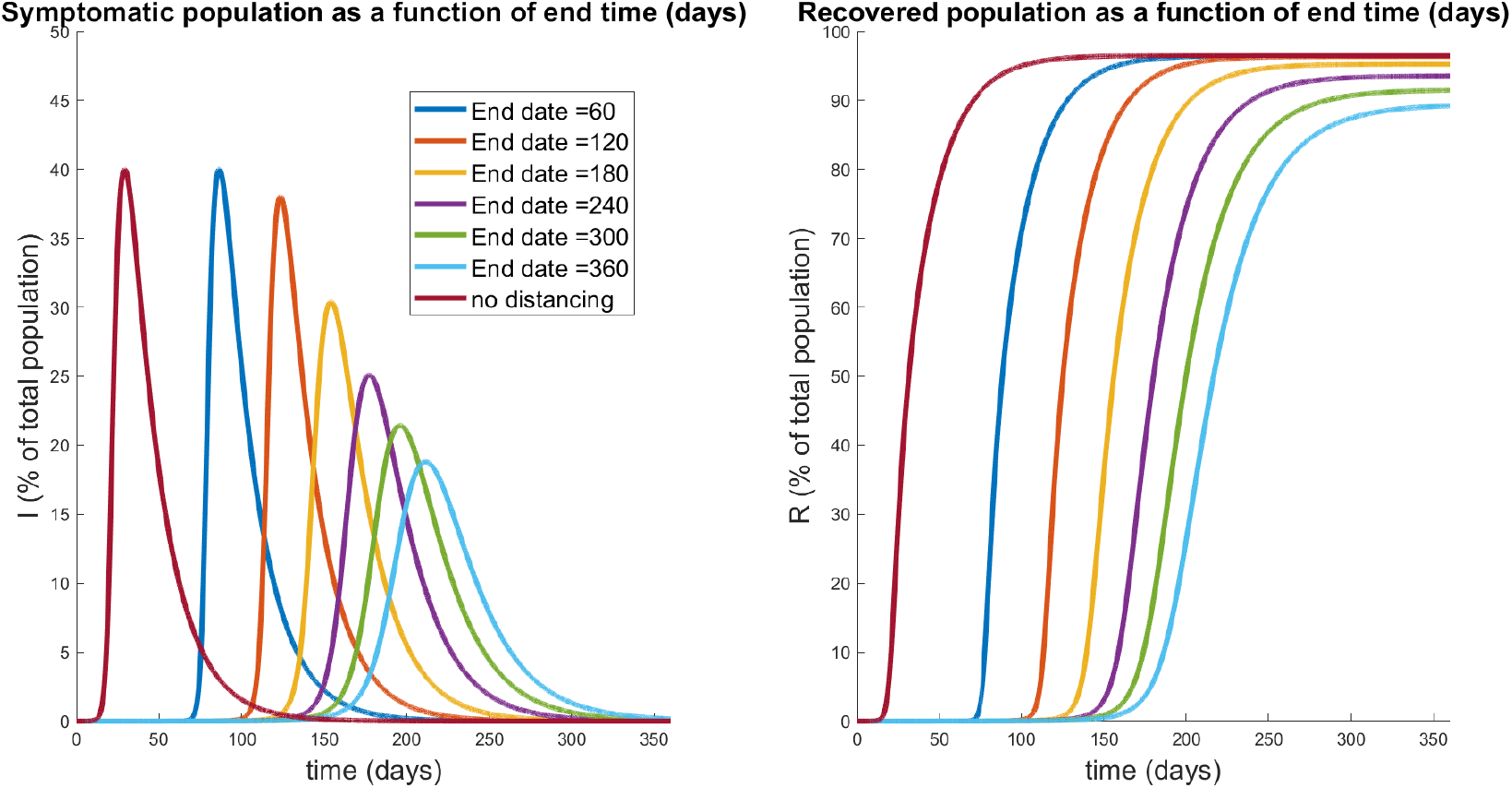
Symptomatic (left) and recovered (right) populations for policies which relax social distancing at a rate determined (inversely) by *t_e_*, the day at which distancing policies are completely removed. The response for no social distancing implemented is included for reference (red curves). Note that *t_e_* = 60 corresponds to immediate relaxation, and has a similar peak to the non-distanced curve. However, gradual relaxation protocols appear to both decrease the peak number of symptomatic individuals, while also spreading out their distribution.

**Figure 18:**
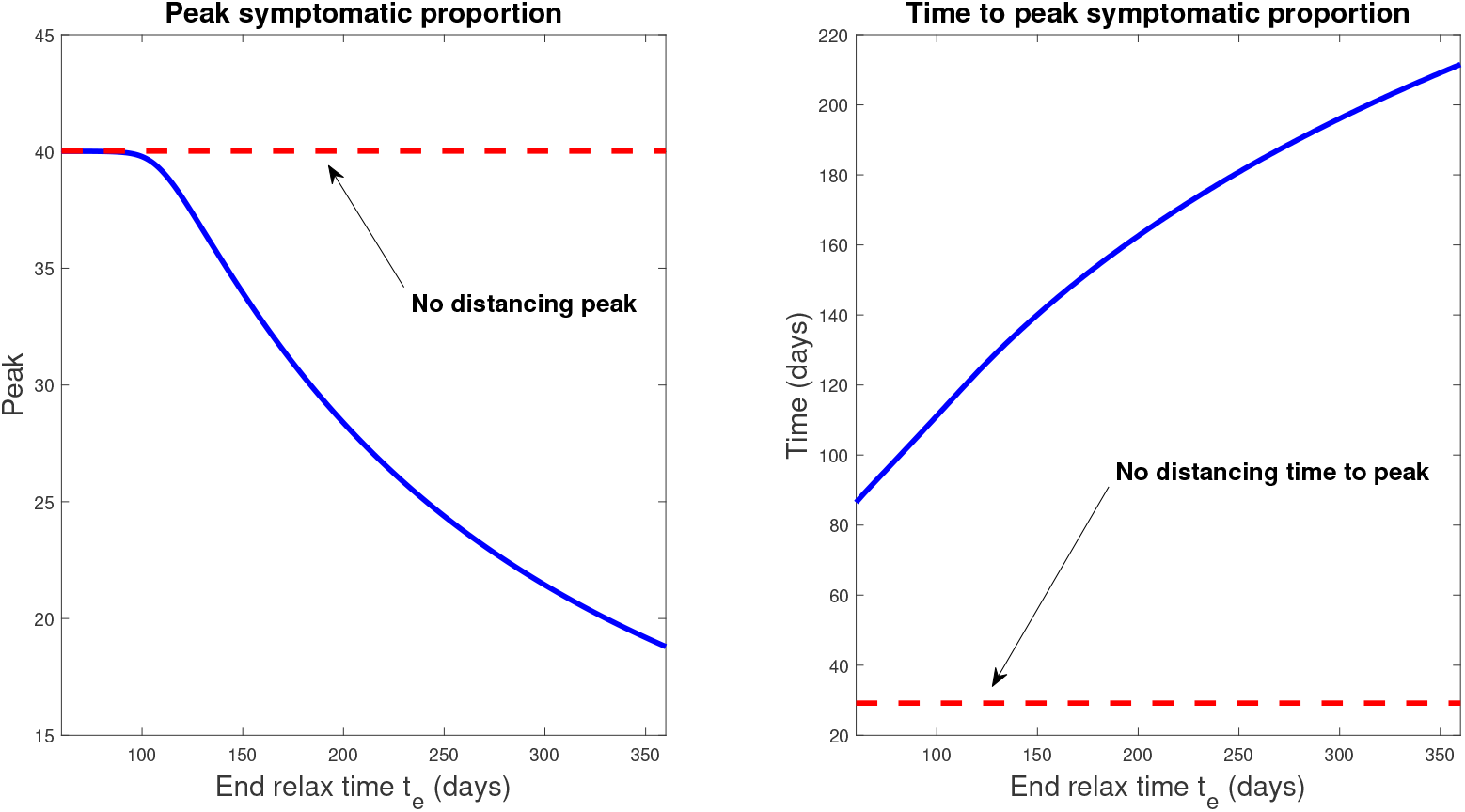
Response of infection dynamics to different relaxation rates after 60 days of social distancing. Policy period is assumed for 360 days. Left panel denotes peak symptomatic population (percentage) at any one time. Right panel is the corresponding time (in days) when this peak occurs. Initial conditions are described in Section 3.2.1.

We also see that, as in Section 3.2.3, all strategies result in a significant fraction of the surviving population being immune by *t_f_* = 360 days; the most gradual relaxation policy has the lowest immune population of just under 90%. In other words, under the assumption that infection confers immunity that lasts at least a year, herd immunity has been largely achieved in all protocols.

#### 3.2.5 Relaxation and a second outbreak

In Section 3.2.4, we saw that the rate of relaxation of social distancing was related to the overall peak symptomatic population: relax too quickly, and the peak is delayed but not inhibited, but gradually lift polices and this peak is both reduced and delayed to a substantial degree. (Figs. 17 and 18). In that analysis, we assumed a fixed distancing period of 60 days, after which relaxation protocols are implemented. In reality however, policies are not designed utilizing artificial timelines, but instead rely on measured data relating to the epidemiology of the outbreak. A question many states and countries now face is the following: once we observe a “flattening of the curve” (e.g. a plateau of new reported cases), how should relaxation be implemented? In this section, we use our model to address this question.

Consider an initial outbreak as discussed in Section 3.2.1 (specifically initial conditions in equation (54)), assuming that an immediate strict social distancing protocol is enforced. Mathematically, this means that

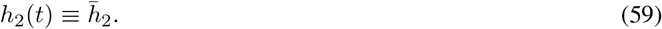

As in the previous sections, we fix

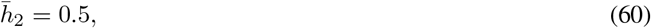

which implies that

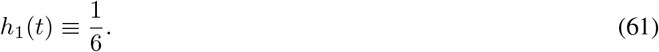

The above policies are applied until the outbreak, here measured as the growth of the symptomatic population, dissipates. That is, we apply the above until a time *t_d_* such that

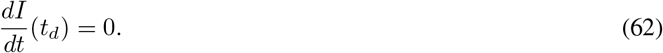

According to our model, once the peak is achieved at time *t_d_*, we see that the “worst is over”. This is confirmed by the decrease in the infective population when extreme social distancing is continued beyond the peak infection time, as shown in Fig. 19(a). Therefore, it is a reasonable policy decision to start to relax social distancing measures once the peak has been obtained. However, the plot of the recovered individuals suggests we need to be careful about how relaxation is implemented. If we completely relaxed all measures immediately at day *t_d_* = 395, only 23% of the population is recovered (and assumed to be immune). According to our calculations of *R*_0_ in Section 3.1 (in particular Fig. 3), *R*_0_ > 1 when *h*_2_ = 0, *R*^*^ = *R*(*t_d_*) = 23% and the number of infectives will increase. This highlights the need to carefully design relaxation policies in order to avoid a second wave. Here, we now explore how to design such a relaxation strategy.

**Figure 19:**
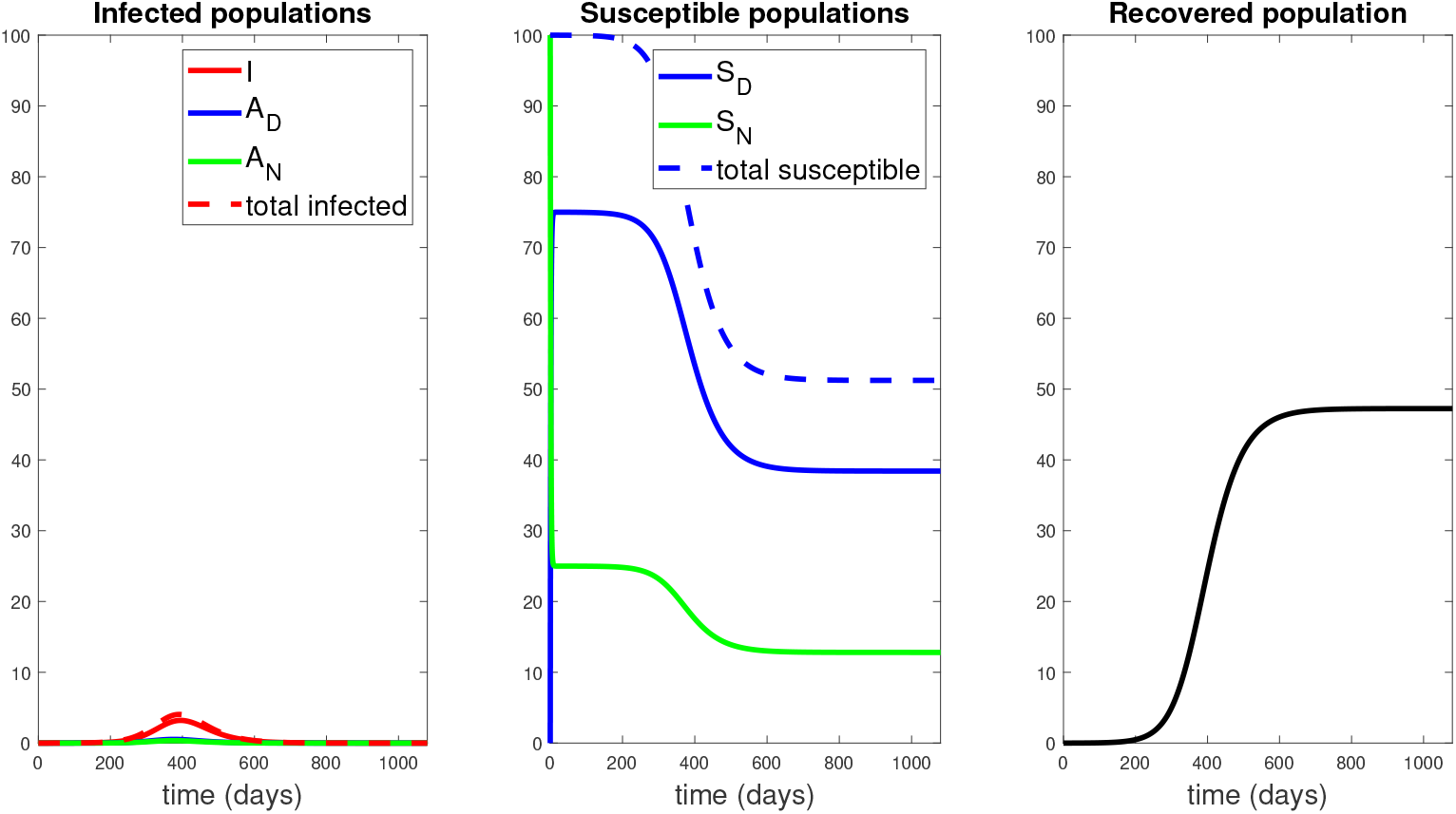
Response to 1080 days of social distancing, with *h*_2_ as in equation (60) for *t* ∈ [0,1080] days. Note the peak of the symptomatic population is approximately 3.2% of the population, occurring at around day 395. As suggested by the right panel, the population is still largely susceptible, and one may suspect that a second epidemic will occur if social distancing protocols are immediately abandoned.

We thus consider a relaxation policy similar to that shown in Fig. 16, with *t_d_* taking the role of *t*_1_, and investigate the response to different relaxation policies (different *t_e_*). From Fig. 19, we see that *t_d_* is given by

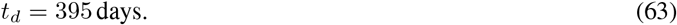

The dynamical response of our model to selected relaxation rates appears in Fig. 20, while a more global characterization is provided in Fig. 21. Note in the left panel of Fig. 20, all protocols have the same response until time *t* = *t_d_* (black curve); this is because social distancing is identically enforced for 0 ≤ *t* ≤ *t_d_*. After *t_d_*, we see a different infection response based upon the rate at which distancing policies are relaxed. Note that if the rate of relaxation is too large (*t_e_* = 400,500,600 days in Fig. 20), we see a second wave of infections, larger than the original peak. For example, if social distancing policies are concluded by day 400 (a very small relaxation period, since relaxing begun on day 395), we see a second peak of symptomatic individuals of over 26% of the population by day 417; compare this with the original peak of 3.2%. However, if relaxation is relatively slow (for example, *t_e_* = 800 days), we see no second wave of infection. Hence in designing policy, we must carefully consider the manner in which social distancing policies are removed; if it is too fast, then we risk undoing the results achieved in the first *t_d_* = 395 days.

**Figure 20:**
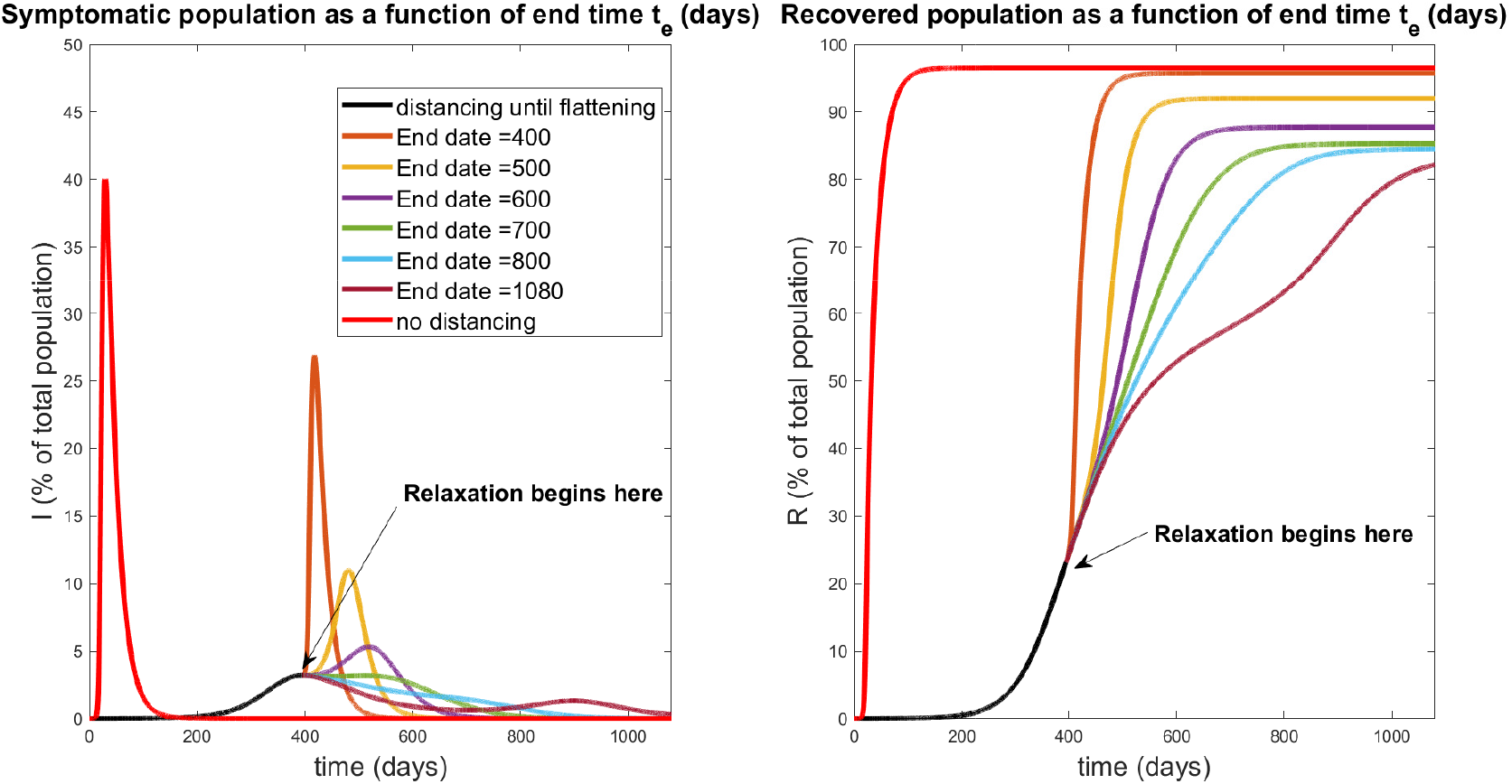
Relaxation that was started during flattening. Symptomatic (left) and recovered (right) populations for policies which relax social distancing at a rate determined (inversely) by *t_e_*, the day at which distancing policies are completely removed. The response for no social distancing implemented is included for reference (red curves). All curves besides the red curve are identical for the first *t_d_* = 395 days of treatment, when social distancing is implemented with 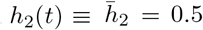 per day for *t* ∈ [0,t_d_]. Note that a second wave occurs for relaxation protocols that end too quickly (*t_e_* = 400, 500, 600 days). However, if relaxation is slow enough (larger *t_e_*), a second wave of infection is completely mitigated. All simulations are taken over 1080 days.

**Figure 21:**
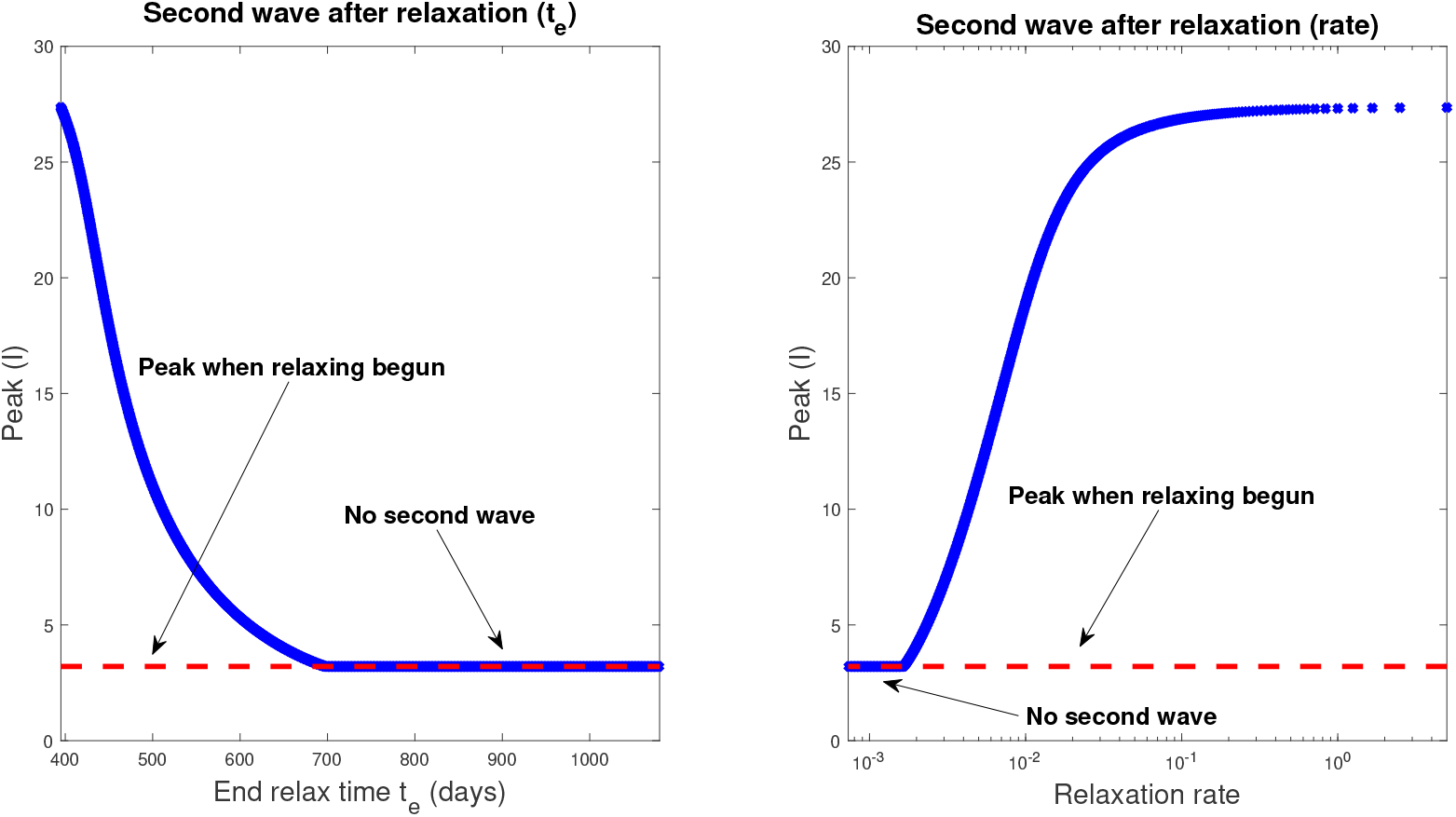
Magnitudes of peak of symptomatic individuals (percentage of total population) as a function of end time *t_e_* (left) and rate (right). Note that *t_e_* and relaxation rate are inversely proportional; see equation (64). Relaxation is begun when the symptomatic population first “flattens,” which is indicated by the dashed red line in the figure, and occurs for all schedules at day *t_d_* = 395. A peak equal to the dashed red line thus indicates that no second wave of infections occurs, i.e. there was never a day when the number of infected individuals was greater than the peak when relaxation was commenced. All simulations are taken over 1080 days.

In Fig. 21, we provide a plot of the peak of the infected population as a function of both the full relaxation time (left) and the relaxation rate (right). Note that the rate is the speed (magnitude) of the relaxation schedule, and corresponds to the absolute value of the slope in Fig. 16. The time *t_e_* and rate are thus related via

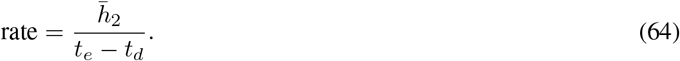

All relaxation policies are initiated at day *t_d_* = 395, where the symptomatic population has reached a peak value of 3.2%. If no second wave occurs (by second wave we mean a peak value of symptomatic individuals *larger* than the original peak at day *t_d_*), then the maximum value of *I* corresponds to this 3.2%, indicated with a dashed red line in both panels of Fig. 20. Hence we compute a critical relaxation rate *r_c_*, such that if social distancing is relaxed faster than *r_c_*, a second outbreak will occur (right panel of Fig. 21). However, if distancing restrictions are eased slowly enough (i.e. slower than *r_c_*), a second peak never occurs, and herd immunity is largely achieved after 1080 days (again, assuming it exists). For our parameters, we find that this critical rate is

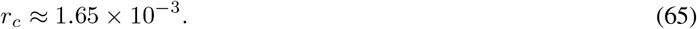

Hence we may provide an estimate of the degree to which social distancing may be relaxed. Of course, this value depends critically on parameter values and other assumptions which remain (as of writing) unknown. Indeed, the main conclusion should not be the exact value given in equation (65), but rather the phenomenon that the “speed” of relaxation has significant consequences for subsequent outbreaks, which we believe is robust with respect to parameter values.

## 4 Discussion & Conclusion

In this work, we have introduced a novel epidemiological model of the COVID-19 pandemic which incorporates explicit social distancing via separate compartments for susceptible and asymptomatic (but infectious) individuals. We believe that this is the first model which characterizes social distancing protocols as rates of flow between these compartments, with the rates determined by guidelines implemented by regional governmental intervention. In particular, we view these rates as controls, and one of our primary focuses of study is disease response (measured by peak symptomatic proportion) to different mandated social distancing controls.

Our major contributions and results are described below:

1. In model formulation, we decouple the rate of social distancing (*h*_2_) with the decrease in contact due to social distancing (*ϵ_S_*, *ϵ_A_*, *ϵ_I_*). The latter we term the *contact rescaling factor (CoRF)*, which should be interpreted as an effectiveness of social distancing. Hence we explicitly account for both the rate at which individuals distance, and how effective distancing is as a means of suppressing viral transmission.
2. The basic reproduction number, *R*_0_, is explicitly calculated for our model system. For parameters obtained from data, we show that at sufficiently early stages of the pandemic, *R*_0_ > 1 unless the rate of social distancing is quite large. This implies that under most realistic circumstances there will be an initial outbreak of the COVID-19 pandemic. However, the situation is not hopeless, as social distancing policies are able to push *R*_0_ < 1 as the disease spreads throughout the population.
3. *R*_0_ is sensitive to the fraction of infective individuals that are asymptomatic, and to the infectivity of these asymptomatic individuals. Hence understanding this population (through, for example, widespread testing) is critical for making informed policy decisions.
4. There is a critical time to implement social distancing guidelines (what we label the *critical implementation delay (CID*)), after which social distancing will have little effect on mitigating the percent of symptomatics at the peak of the outbreak. Surprising, the CID occurs well before the peak symptomatic proportion would have originally appeared under non-distanced protocols (CID is approximately two weeks for our parameter values, while the non-distanced peak occurs at about 30 days).
5. While implementing distancing faster than the CID does not significantly change the number infected at the peak, it does significantly alter *when* this peak occurs. For example, implementing social distancing at day 5 instead of day 15 pushes the peak forward by nearly a year, which allows time for the development of a therapeutic.
6. Periodic relaxation strategies, where normal behavior is allowed for certain periods of time, can significantly reduce the symptomatic burden. However, such scheduling is not robust, and small errors (either in timing, or via parameter estimation) may have catastrophic repercussions.
7. Gradual relaxation can substantially improve the overall symptomatic response, but the rate of relaxation is important to prevent a “second wave” of virus outbreak. Prolonged relaxation, or sufficiently slow relaxation upon flattening, can significantly “flatten the curve.”

As noted throughout the manuscript, exact predictions rely on estimated parameter values, which currently vary widely throughout the literature. On the other hand, we believe that the qualitative phenomenon observed are robust, and should be considered during policy design.

There are several directions for future work. First, our model has shown that the effectiveness of social distancing policies is sensitively dependent on when measures are implemented. By calibrating the model to the dynamics of a particular locale, we could determine a threshold case load at which the population is at high risk of a second wave of infection. Knowledge of this threshold could help policy makers determine if/when social distancing recommendations need to be strengthened. Second, we will conduct a more systematic control analysis involving both the rate of social distancing (*h*_2_) and the stringency of distancing (CoRF, i.e. *ϵ* terms). Ideally, we would like to minimize the peak of the symptomatic population (*I*) while simultaneously maximizing the time to reach this peak; we view such an objective as a precise quantification of “flattening the curve.” This must be done with distancing constraints imposed, to reflect the fact that a certain percentage of the population must remain active to maintain a functional society (healthcare workers, food supply, emergency responders, etc.). Using optimal control and feedback laws, our model can help inform policy makers as they make difficult decisions about how to adapt to the ongoing pandemic.

## Data Availability

The data used is from public sources.

## Author Contributions

Conception and design: All authors.

Development of methodology: All authors.

Analysis and interpretation of data: All authors.

Writing, review and/or revision of manuscript: All authors.

## A The Basic Reproduction Number *R*_0_

The next generation matrix algorithm, proposed by Diekmann at. al. in 1990 [40], is a technique used to calculate the basic reproduction number *R*_0_. We explain it briefly, for more details see [41, 43].

Here are the steps to compute the next generation matrix *G*:

1. Let X = {*x*_1_, *x*_2_,…, *x_n_*} represent the *n* infected host compartments, and *Y* = {*y*_1_, *y*_2_,…, *y_m_*} represent the *m* other host compartments.
2. Write your ODE system as:

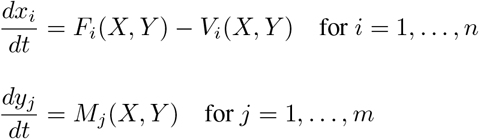

where *F_i_* represents the rate at which new infectives enter compartment *i*, and *V_i_* represents the transfer of individuals out of and into the *i*-th compartment.
3. Let *F_X_* and *V_X_* represent the Jacobian matrices evaluated at the DFSS of the vector fields

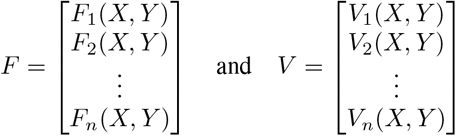

respectively.
4. The next generation matrix G is defined by

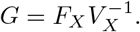 *G* is a non-negative matrix with an eigenvalue which is real, positive, and strictly greater than all the others. This largest eigenvalue is *R*_0_.

### A.1 *R*_0_ for the six-compartment SIR model (equations (8)-(13))

We compute the basic reproduction number *R*_0_ of the six-compartment SIR model with the Next Generation Matrix Algorithm [40,41, 43].

Let

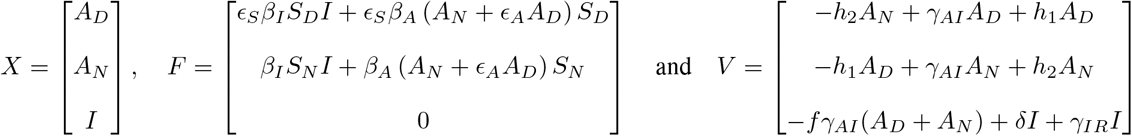

Thus, *Ẋ* = *F* − *V* and

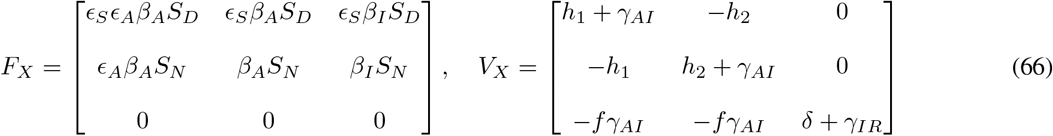

where *F_X_* and *V_X_* denote the Jacobian matrices of F and V respectively; and

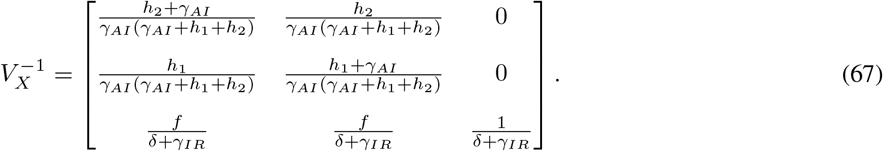

Let *g_ij_* represent the (*i*, *j*) –entry of the next generation matrix *G*. At the DFSS (where we must satisfy the equation 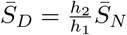) we have that,

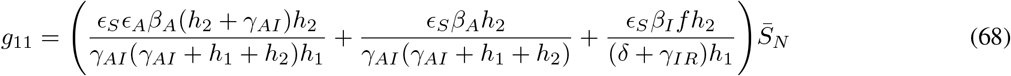

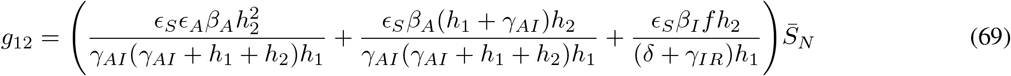

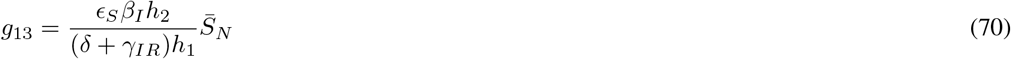

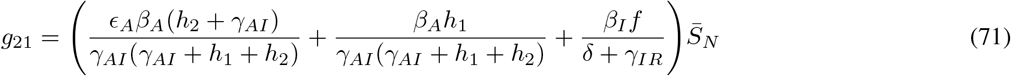

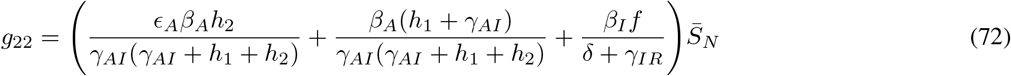

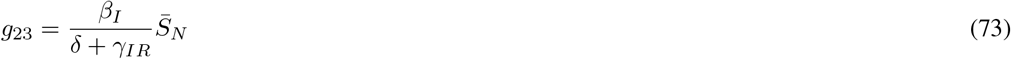

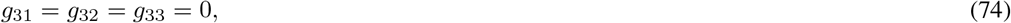

where 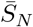 represents the value of *S_N_* at DFSS. The characteristic polynomial of *G* is:

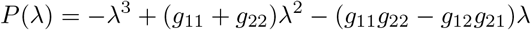

with roots (eigenvalues of *G*):

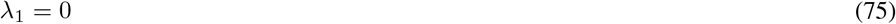

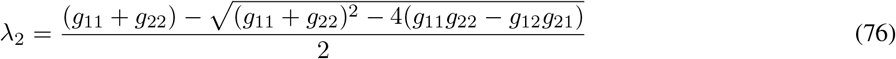

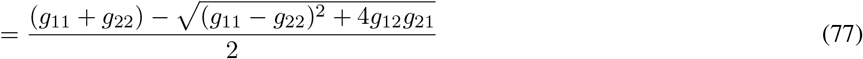

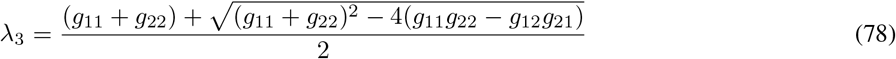

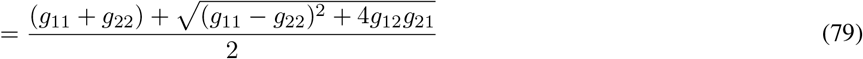

Therefore, λ_3_ is the basic reproduction number, *R*_0_.

#### A.1.1 Conditions for *R*_0_ < 1. The case *h*_2_ =0 for the six-compartment SIR model

If *h*_2_, the rate of social distancing, is equals to zero (which implies 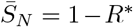). The basic reproduction number reduces to:

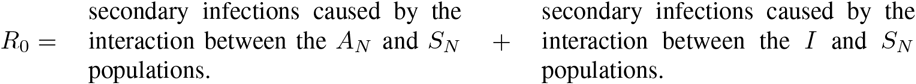

In other words,

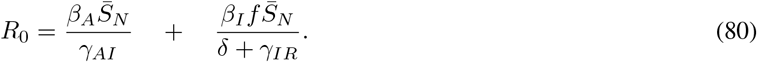

For *R*_0_ to be less than 1 we must satisfy the equation:

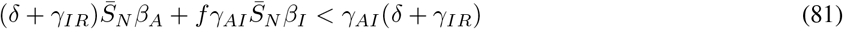

or equivalently,

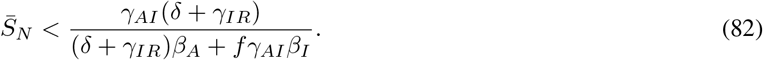

#### A.1.2 Conditions for *R*_0_ < 1. The general case for the six-compartment SIR model

Let 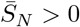, and consider the polynomial

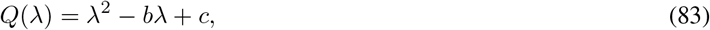

where *b* and *c* are the trace and determinant of the matrix

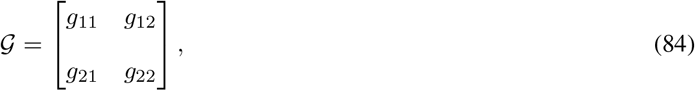

Remark 1.

1. 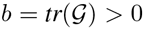
2. *The discriminant* 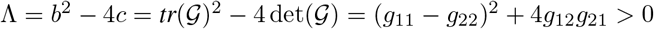.
3. *The eigenvalues of* 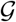 *are:*

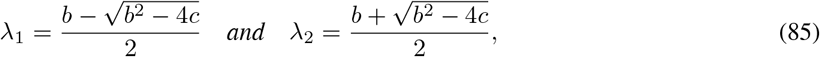 *where* λ_1_ < λ_2_ *and* λ_2_ > 0.
4. *Let*

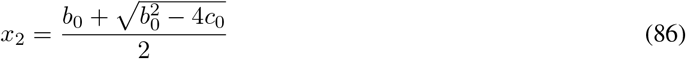

*where*

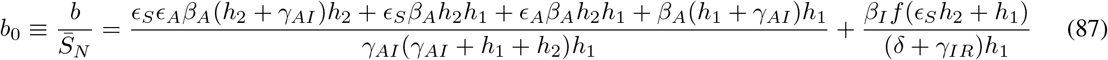

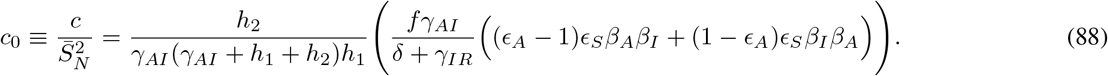 Thus, 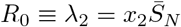.
5. At the *disease-free equilibrium, the following equations are satisfied:*

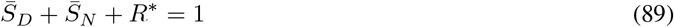

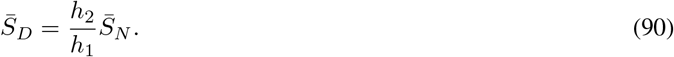

*which gives the relation*

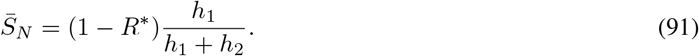 *Therefore*,

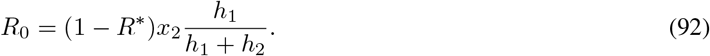
6. From *equation* (92), *it follows that R*_0_ < 1 if and only if

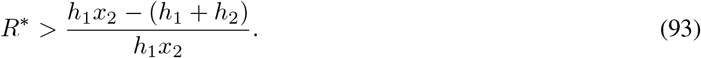
7. For 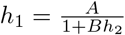, *equation* (93) is *equivalent to*

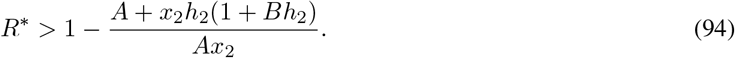

### A.2 *R*_0_ for the seven-compartment SIR model (equations (1)-(7))

Let

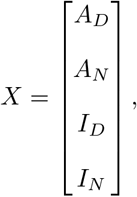

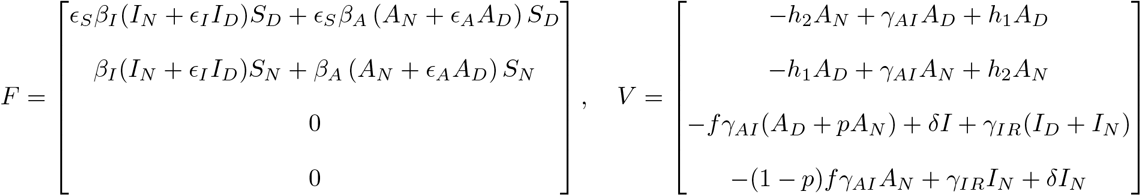

Thus, *Ẋ* = *F* − *V* and

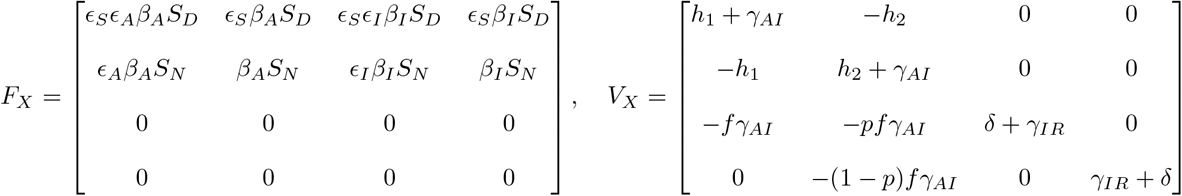

where *F_X_* and *V_X_* denote the Jacobian matrices of *F* and V respectively; and

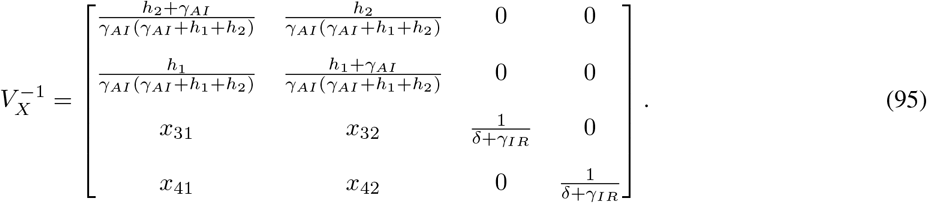

with

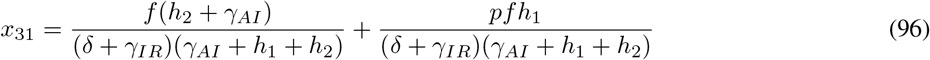

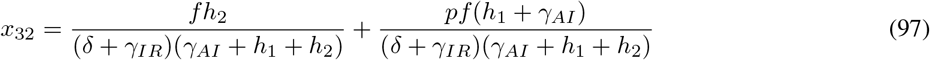

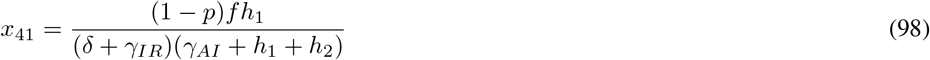

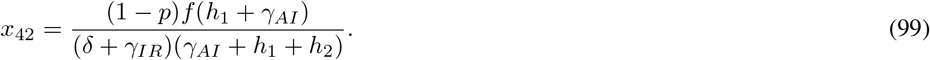

is the inverse matrix of *V_X_*.

Let *g_ij_* represents the (*i*, *j*) –entry of the next generation matrix *G*. At the DFSS (where we must satisfy the equation 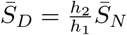) we have that,

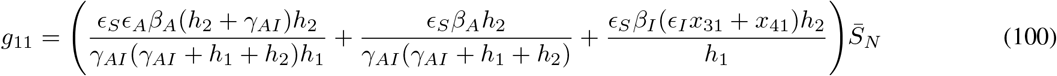

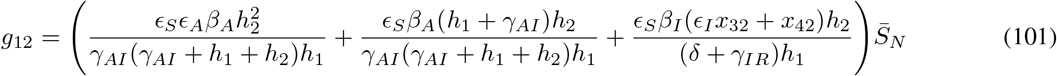

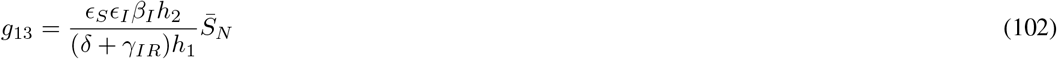

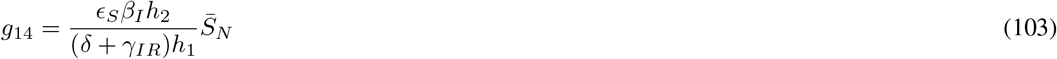

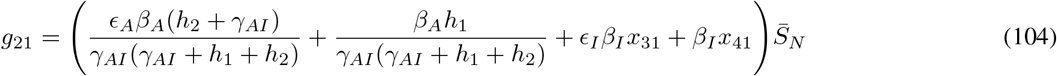

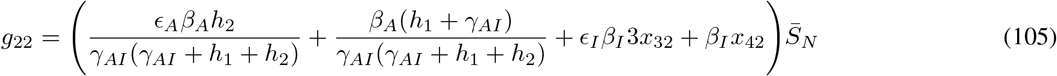

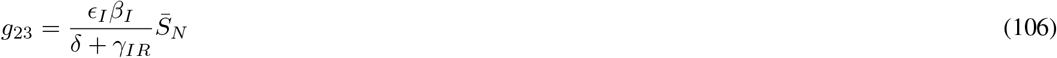

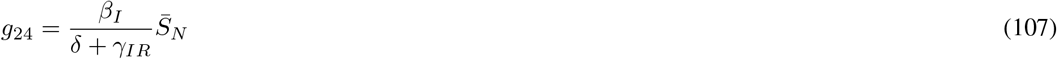

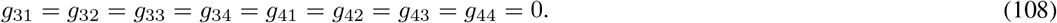

Thus,

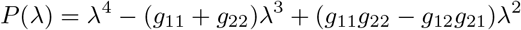

is the characteristic polynomial of *G* with roots:

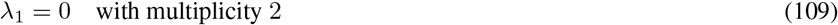

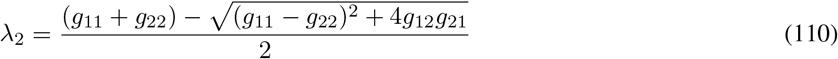

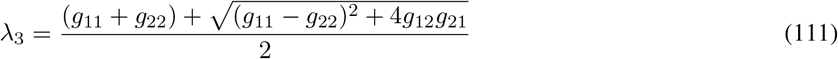

Therefore, λ_3_ is the basic reproduction number, *R*_0_.

## B Heatmaps Corresponding to Contour Plots of *R*_0_

**Figure 22:**
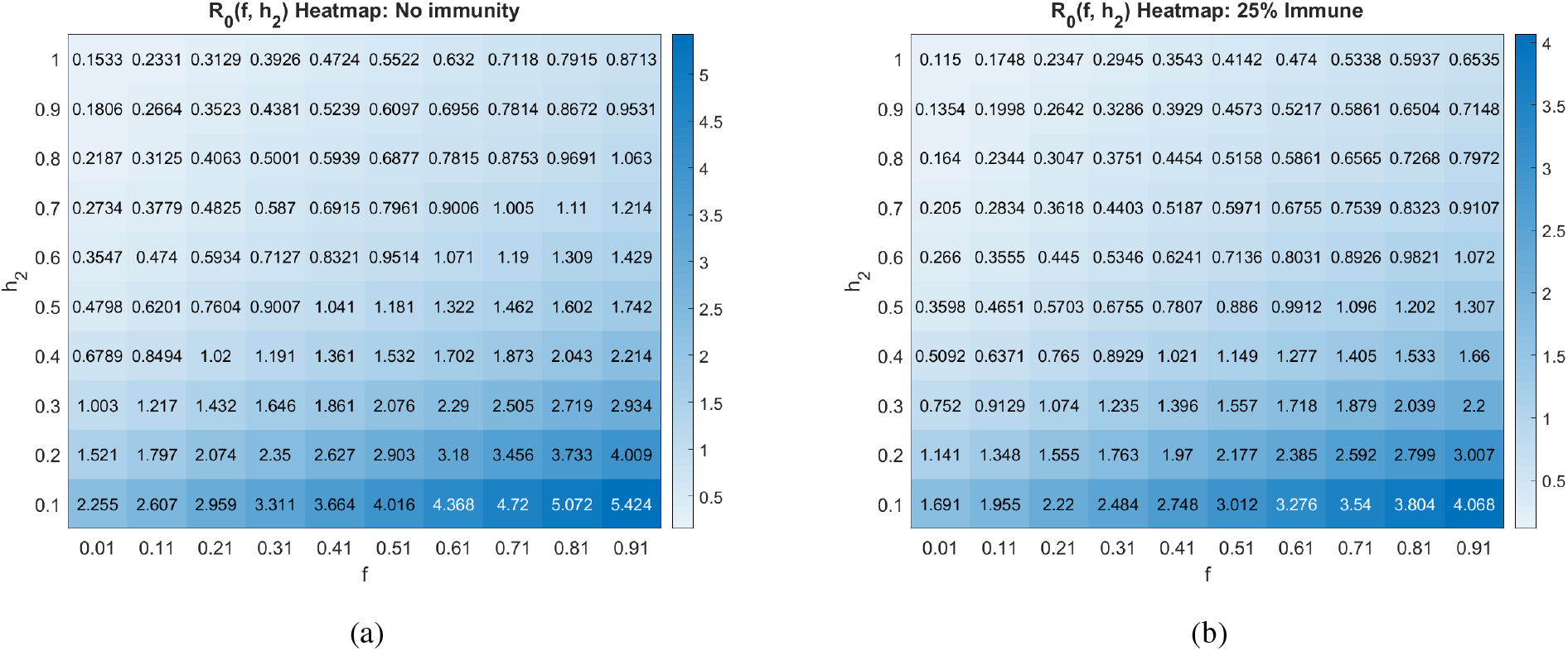
Basic reproduction number as a function of the social distancing rate parameter *h*_2_ and fraction of individuals who become symptomatic (*f*) at different pandemic stages. All other parameters as in Table 2.

**Figure 23:**
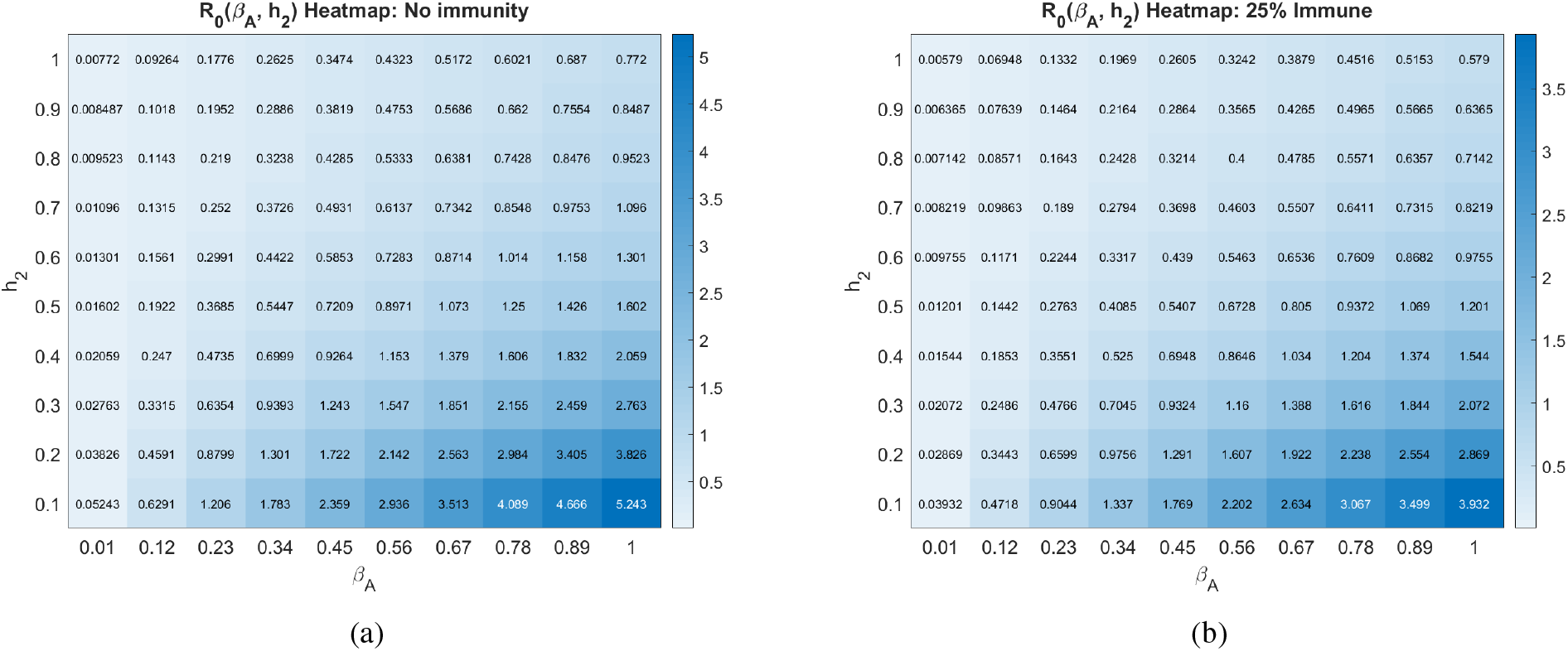
Basic reproduction number as a function of the social distancing rate parameter *h*_2_ and infectivity rate of asymptomatics *β_A_* at different pandemic stages. All other parameters as in Table 2.

**Figure 24:**
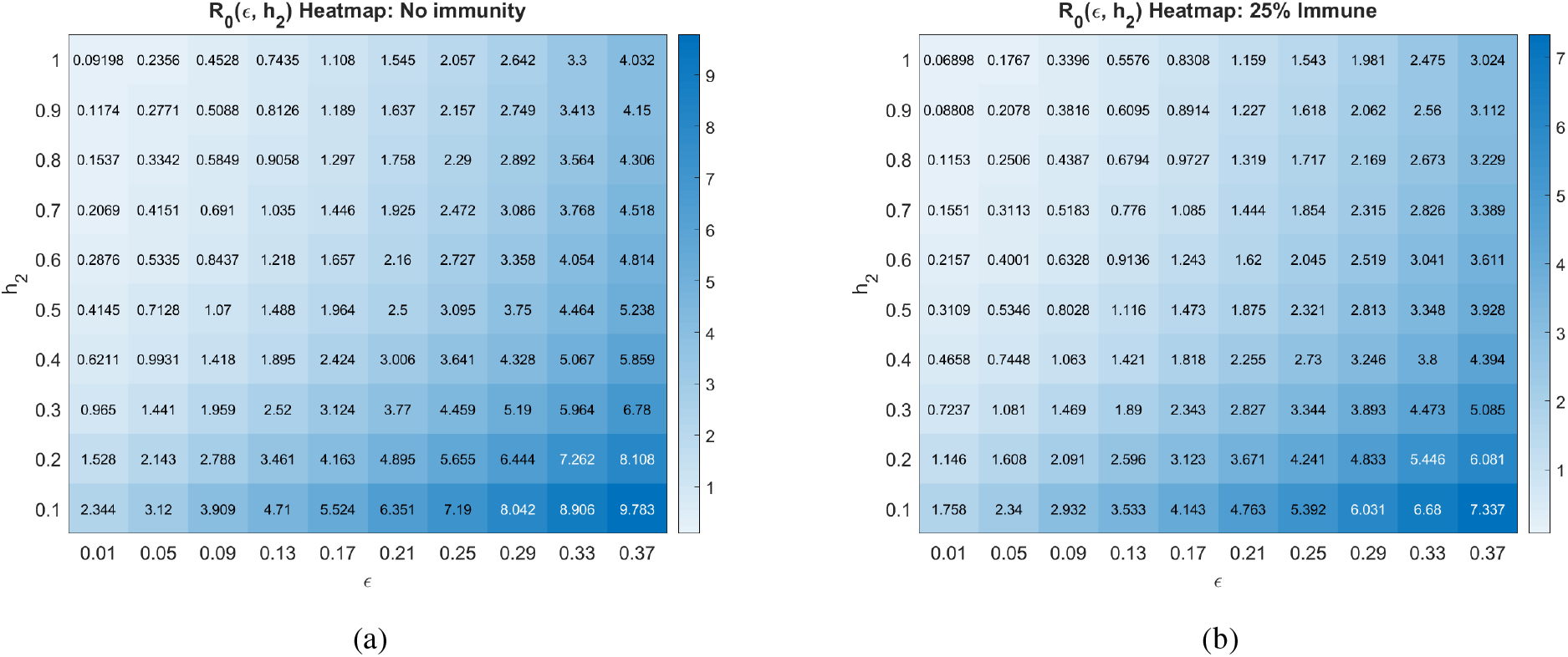
Basic reproduction number as a function of the social distancing rate parameter *h*_2_ and the contact rescaling factor (CoRF) *ϵ* at different pandemic stages. CoRF measures the impact of social distancing on infectivity rate. All other parameters as in Table 2.

## C Convergence of infectives to zero

It is a routine exercise to show that, in our model, *I*(*t*) → 0 as *t* → ∞, and similarly for *A_N_* (*t*) and *A_D_* (*t*). Indeed, consider the total population fraction *N* defined by *N* := *S_D_* +*S_N_*+*A_D_* +*A_N_*+*I*+*R* and observe that *dN*/*dt* = −*δI* ≤ 0. The function *N* is continuously differentiable. The LaSalle Invariance Principle [44] implies that all solutions converge to an invariant set Ω included in *dN*/*dt* = 0, meaning in particular that all solutions have *I*(*t*) → 0, as claimed. Furthermore, the equation *dI*/*dt* = *fγ_AI_*(*A_D_* + *A_N_*) − *δI* − *γ_IR_I* when restricted to Ω says that 0 = *fγ_AI_*(*A_D_*(*t*) + *A_N_*(*t*)), which means that, in this set to which all solutions converge, both *A_D_* and *A_N_* are identically zero.

As *dR*/*dt* = 0 on this set Ω, *R*(*t*) converges to a limit *r* (which is in general nonzero). The equations for susceptibles become, in Ω:

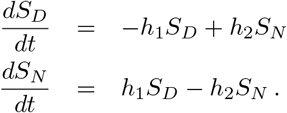

Thus *S_D_* and *S_N_* equilibrate to constant values under the constraint that *S_D_* + *S_N_* = *n* − *r*, where *n* =lim_t→∞_*N*(*t*), i.e. 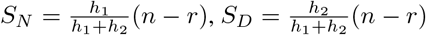.

## D Approximation of 6 compartment model as an *SAIR* system

Here, we remark that, in some cases, the model introduced in Section 2.1 can be approximated by a four-compartment *SAIR* (Susceptible-Asymptomatic-Infected-Recovered) model with combined socially distanced and non-distanced populations. Specifically, we proceed under the assumption that there exists a time-scale separation, so that the social distancing dynamics occur much faster than dynamics governed by infection-specific parameters. That is, at any given instant of time *t*, we assume that distancing instantly calibrates to its equilibrium distribution, i.e. that

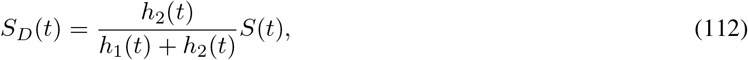

where *S*(*t*) is the total susceptible population:

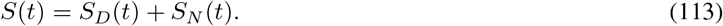

Denoting the fraction in equation (112) as,

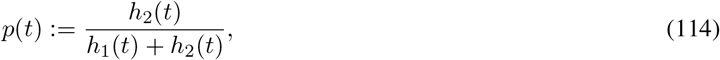

we thus have that at any time *t*,

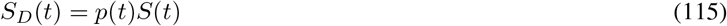

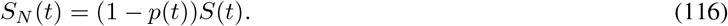

Similarly,

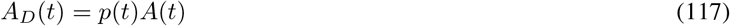

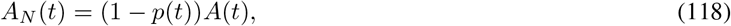

where

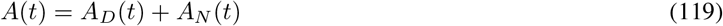

is the total asymptomatic population. The above assumption thus allows us to convert to a four-compartment *SAIR* model, with *S* and *A* given by the *total* susceptible and asymptomatic populations, respectively. For instance, we can calculate 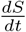 as

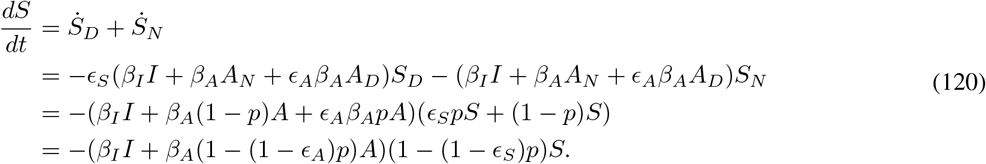

Note that the above takes the form of a more standard *SEIR* model, with *A* = *E* and the *E* compartment being infectious. Recall that the control appears in *p*(*t*):

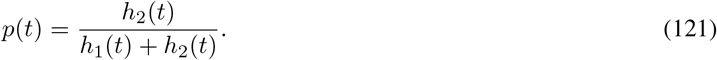

However, in the *Ṡ* equation above, the *p* term is simply altering the transmission rates *β* (decreasing *β_I_* and *β_A_* by increasing *h*_2_). Hence we see that manipulating *h*_2_ (as we do in this manuscript) is analogous to manipulating *β* (as is often done in recent works related to controlling COVID-19), at least in the case where a fast response to distancing mandates is assumed. The derivation makes explicit how the *β* depends on the rates of distancing.

For completeness, the simplified *SAIR* model takes the below form (calculations for other compartments are similar):

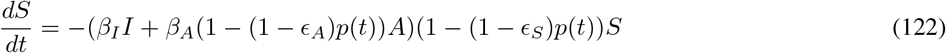

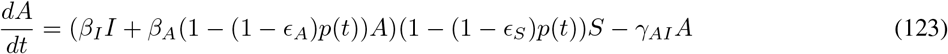

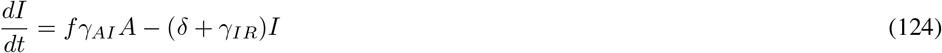

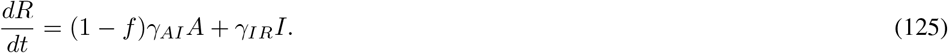

## Footnotes

1 The use of the letter “*R*” for “recovered” and for “*R*_0_” is an unfortunate coincidence.

## References

[1] D. Adam. Special report: The simulations driving the world’s response to COVID-19. Nature, 580(7803):316–318, 04 2020.

[2] N. Ferguson, D. Laydon, G. Nedjati-Gilani, N. Imai, K. Ainslie, M. Baguelin, S. Bhatia, A. Boonyasiri, Z. Cucunuba, G. Cuomo-Dannenburg, et al. Impact of non-pharmaceutical interventions (npis) to reduce COVID-19 mortality and healthcare demand. Imperial College, London. DOI: https://doi.org/10.25561/77482, 2020.

[3] J. Lourenco, R. Paton, M. Ghafari, M. Kraemer, C. Thompson, P. Simmonds, P. Klenerman, and S. Gupta. Fundamental principles of epidemic spread highlight the immediate need for large-scale serological surveys to assess the stage of the sars-cov-2 epidemic. *medRxiv*, 2020.

[4] C. Murray. Forecasting the impact of the first wave of the covid-19 pandemic on hospital demand and deaths for the usa and european economic area countries. *medRxiv*, 2020.

[5] X. Liu, G. Hewings, S. Wang, M. Qin, X. Xiang, S. Zheng, and X. Li. Modeling the situation of covid-19 and effects of different containment strategies in china with dynamic differential equations and parameters estimation. *medRxiv*, 2020.

[6] M. Chinazzi, J. Davis, M. Ajelli, C. Gioannini, M. Litvinova, S. Merler, A. Pastore y Piontti, K. Mu, L. Rossi, K. Sun, C. Viboud, X. Xiong, H. Yu, M. Halloran, I. Longini, and A. Vespignani. The effect of travel restrictions on the spread of the 2019 novel coronavirus (covid-19) outbreak. Science, 368(6489):395–400, 2020.

[7] H. Tian, Y. Liu, Y. Li, C. Wu, B. Chen, M. Kraemer, B. Li, J. Cai, B. Xu, Q. Yang, B. Wang, P. Yang, Y. Cui, Y. Song, P. Zheng, Q. Wang, O. Bjornstad, R. Yang, B. Grenfell, O. Pybus, and C. Dye. The impact of transmission control measures during the first 50 days of the covid-19 epidemic in china. *medRxiv*, 2020.

[8] A. Kucharski, T. Russell, C. Diamond, Y. Liu, J. Edmunds, S. Funk, R. Eggo, F. Sun, M. Jit, J. Munday, N. Davies, A. Gimma, K. Van Zandvoort, H. Gibbs, J. Hellewell, C. Jarvis, S. Clifford, B. Quilty, N. Bosse, S. Abbott, P. Klepac, and S. Flasche. Early dynamics of transmission and control of COVID-19: a mathematical modelling study. Lancet Infect Dis, 20(5):553–558, 05 2020.

[9] S. Park, D. Cornforth, J. Dushoff, and J. Weitz. The time scale of asymptomatic transmission affects estimates of epidemic potential in the covid-19 outbreak. *medRxiv*, 2020.

[10] F. Brauer. Some simple epidemic models. Mathematical Biosciences and Engineering, 3(1), 2006.

[11] H. Hethcote, M. Zhien, and L. Shengbing. Effects of quarantine in six endemic models for infectious diseases. Mathematical Biosciences, 180, 2002.

[12] G. Giordano, F. Blanchini, R. Bruno, P. Colaneri, A. Di Filippo, A. Di Matteo, and M. Colaneri. Modelling the COVID-19 epidemic and implementation of population-wide interventions in Italy. Nat. Med., Apr 2020.

[13] F. Casella. Can the covid-19 epidemic be managed on the basis of daily data? *arXiv preprint arXiv:2003.06967*, 2020.

[14] F. Di Lauro, I. Kiss, and J. Miller. The timing of one-shot interventions for epidemic control. *medRxiv*, 2020.

[15] D. Morris, F. Rossine, J. Plotkin, and S. Levin. Optimal, near-optimal, and robust epidemic control. OSF preprint, 2020.

[16] M. Bin, P. Cheung, E. Crisostomi, P. Ferraro, H. Lhachemi, R. Murray-Smith, C. Myant, T. Parisini, R. Shorten, S. Stein, and L. Stone. On fast multi-shot covid-19 interventions for post lock-down mitigation, 2020.

[17] W. Kermack and A. McKendrick. A contribution to the mathematical theory of epidemics. The Royal Society Publishing, 114(772), 1927.

[18] X. Liu and P. Stechlinski. Infectious Disease Modeling, volume 19. Springer Nature, Cham, 2017.

[19] H. Mo, G. Zeng, X. Ren, H. Li, C. Ke, Y. Tan, C. Cai, K. Lai, R. Chen, M. Chan-Yeung, and N. Zhong. Longitudinal profile of antibodies against sars-coronavirus in sars patients and their clinical significance. Respirology, 11, 2006.

[20] L. Wu, N. Wang, Y. Chang, X. Tian, D. Na, L. Zhang, L. Zheng, T. Lan, L. Wang, and G. Liang. Duration of antibody responses after severe acute respiratory syndrome. Emerg Infect Dis., 13(10), 2007.

[21] W. Cao, W. Liu, P. Zhang, F. Zhang, and J. Richardus. Disappearance of antibodies to sars-associated coronavirus after recovery. New England Journal of Medicine, 357(11):1162–1163, 2007.

[22] S. Lauer, K. Grantz, Q. Bi, F. Jones, Q. Zheng, H. Meredith, A. Azman, N. Reich, and J. Lessler. The Incubation Period of Coronavirus Disease 2019 (COVID-19) From Publicly Reported Confirmed Cases: Estimation and Application. Annals of Internal Medicine, 172(9):577–582, 05 2020.

[23] Q. Li, X. Guan, P. Wu, X. Wang, L. Zhou, Y. Tong, R. Ren, K. Leung, E. Lau, J. Wong, X. Xing, N. Xiang, Y. Wu, C. Li, Q. Chen, D. Li, T. Liu, J. Zhao, M. Liu, W. Tu, C. Chen, L. Jin, R. Yang, Q. Wang, S. Zhou, R. Wang, H. Liu, Y. Luo, Y. Liu, G. Shao, H. Li, Z. Tao, Y. Yang, Z. Deng, B. Liu, Z. Ma, Y. Zhang, G. Shi, T. Lam, J. Wu, G. Gao, B. Cowling, B. Yang, G. Leung, and Z. Feng. Early Transmission Dynamics in Wuhan, China, of Novel Coronavirus-Infected Pneumonia. N. Engl. J. Med., 382(13):1199–1207, 03 2020.

[24] S. Lauer, K. Grantz, Q. Bi, F. Jones, Q. Zheng, H. Meredith, A. Azman, N. Reich, and J. Lessler. The incubation period of coronavirus disease 2019 (covid-19) from publicly reported confirmed cases: estimation and application. Annals of internal medicine, 2020.

[25] WHO-China Joint Mission. Report of the WHO-China Joint Mission on Coronavirus Disease 2019 (COVID-19). geneva 2020, 2020.

[26] K. Mizumoto, K. Kagaya, A. Zarebski, and G. Chowell. Estimating the asymptomatic proportion of coronavirus disease 2019 (COVID-19) cases on board the diamond princess cruise ship, yokohama, japan, 2020. Eurosurveillance, 25(10), 2020.

[27] M. Day. Covid-19: identifying and isolating asymptomatic people helped eliminate virus in italian village. Bmj, 368:m1165, 2020.

[28] J. Al-Tawfiq. Asymptomatic coronavirus infection: Mers-cov and sars-cov-2 (covid-19). Travel Med Infect Dis, 101608, 2020.

[29] X. Zhou, Y. Li, T. Li, and W. Zhang. Follow-up of asymptomatic patients with sars-cov-2 infection. Clinical Microbiology and Infection, 2020.

[30] H. Nishiura, T. Kobayashi, T. Miyama, A. Suzuki, S. Jung, K. Hayashi, R. Kinoshita, Y. Yang, B. Yuan, A. Akhmetzhanov, et al. Estimation of the asymptomatic ratio of novel coronavirus infections (covid-19). *medRxiv*, 2020.

[31] A. Kimball. Asymptomatic and presymptomatic sars-cov-2 infections in residents of a long-term care skilled nursing facility—king county, washington, march 2020. MMWR. Morbidity and mortality weekly report, 69, 2020.

[32] B. Quilty, S. Clifford, S. Flasche, R. Eggo, et al. Effectiveness of airport screening at detecting travellers infected with novel coronavirus (2019-ncov). Eurosurveillance, 25(5), 2020.

[33] S. Tian, N. Hu, J. Lou, K. Chen, X. Kang, Z. Xiang, H. Chen, D. Wang, N. Liu, D. Liu, et al. Characteristics of covid-19 infection in beijing. Journal of Infection, 2020.

[34] W. Sun, F. Ling, J. Pan, J. Cai, Z. Miao, S. Liu, W. Cheng, and E. Chen. Epidemiological characteristics of 2019 novel coronavirus family clustering in zhejiang province. Zhonghua yu Fang yi xue za zhi [Chinese Journal of Preventive Medicine], 54:E027–E027, 2020.

[35] E. Lavezzo, E. Franchin, C. Ciavarella, G. Cuomo-Dannenburg, L. Barzon, C. Del Vecchio, L. Rossi, R. Manganelli, A. Loregian, N. Navarin, et al. Suppression of covid-19 outbreak in the municipality of vo, italy. *medRxiv*, 2020.

[36] COVID-19 pandemic planning scenarios. https://www.cdc.gov/coronavirus/2019-ncov/hcp/planning-scenarios.html. Accessed: 2020-05-26.

[37] E. Dong, H. Du, and L. Gardner. An interactive web-based dashboard to track covid-19 in real time. The Lancet infectious diseases, 2020.

[38] COVID-Lab: Mapping COVID-19 in your community. https://policylab.chop.edu/covid-lab-mapping-covid-19-your-community. Accessed: 2020-05-26.

[39] R. Li, S. Pei, B. Chen, Y. Song, T. Zhang, W. Yang, and J. Shaman. Substantial undocumented infection facilitates the rapid dissemination of novel coronavirus (sars-cov2). Science, 2020.

[40] O. Diekmann, J. Heesterbeek, and J. Metz. On the definition and the computation of the basic reproduction ratio *R*_0_ in models for infectious diseases in heterogeneous populations. J. Math. Biol., 28, 1990.

[41] O. Diekmann, J. Heesterbeek, and M. Roberts. The construction of next-generation matrices for compartmental epidemic models. J. R. Soc. Interface. DOI: 10.1098/rsif.2009.0386, 7, 2010.

[42] H. Yao, X. Lu, Q. Chen, K. Xu, Y. Chen, L. Cheng, F. Liu, Z. Wu, H. Wu, C. Jin, et al. Patient-derived mutations impact pathogenicity of sars-cov-2. CELL-D-20-01124, 2020.

[43] F. Brauer and C. Castillo-Chavez. Mathematical Models in Population Biology and Epidemiology. Springer, USA, 2010. Second Edition.

[44] H. Khalil. Nonlinear Systems, Third Edition. Prentice Hall, Upper Saddle River, NJ, 2002.

